# Rare variant association analysis in 51,256 type 2 diabetes cases and 370,487 controls informs the spectrum of pathogenicity of monogenic diabetes genes

**DOI:** 10.1101/2023.09.28.23296244

**Authors:** Philip Schroeder, Ravi Mandla, Alicia Huerta-Chagoya, Ahmed Alkanak, Dorka Nagy, Lukasz Szczerbinski, Jesper G.S. Madsen, Joanne B. Cole, Bianca Porneala, Kenneth Westerman, Josephine H. Li, Toni I. Pollin, Jose C. Florez, Anna L. Gloyn, Inês Cebola, Alisa Manning, Aaron Leong, Miriam Udler, Josep M. Mercader

**Author notes:** These authors equally contributed to this work (Philip Schroeder, Ravi Mandla, Alicia Huerta-Chagoya). These authors jointly directed this work (Alisa Manning, Aaron Leong, Miriam Udler, Josep M. Mercader). **Corresponding author:** Josep M Mercader, Programs in Metabolism and Medical and Population Genetics, Broad Institute of Harvard and MIT, 75 Ames St, 02142, Cambridge, MA, United States of America.

## Abstract

We meta-analyzed array data imputed with the TOPMed reference panel and whole-genome sequence (WGS) datasets and performed the largest, rare variant (minor allele frequency as low as 5×10^−5^) GWAS meta-analysis of type 2 diabetes (T2D) comprising 51,256 cases and 370,487 controls.

We identified 52 novel variants at genome-wide significance (*p*<5 × 10^−8^), including 8 novel variants that were either rare or ancestry-specific. Among them, we identified a rare missense variant in *HNF4A* p.Arg114Trp (OR=8.2, 95% confidence interval [CI]=4.6-14.0, *p* = 1.08×10^−13^), previously reported as a variant implicated in Maturity Onset Diabetes of the Young (MODY) with incomplete penetrance. We demonstrated that the diabetes risk in carriers of this variant was modulated by a T2D common variant polygenic risk score (cvPRS) (carriers in the top PRS tertile [OR=18.3, 95%CI=7.2-46.9, *p*=1.2×10^−9^] vs carriers in the bottom PRS tertile [OR=2.6, 95% CI=0.97-7.09, *p* = 0.06]. Association results identified eight variants of intermediate penetrance (OR>5) in monogenic diabetes (MD), which in aggregate as a rare variant PRS were associated with T2D in an independent WGS dataset (OR=4.7, 95% CI=1.86-11.77], *p* = 0.001). Our data also provided support evidence for 21% of the variants reported in ClinVar in these MD genes as benign based on lack of association with T2D.

Our work provides a framework for using rare variant imputation and WGS analyses in large-scale population-based association studies to identify large-effect rare variants and provide evidence for informing variant pathogenicity.

## Introduction

Large genome-wide association meta-analyses have allowed the discovery of hundreds of genetic variants, mostly common (minor allele frequency [MAF] > 0.05), associated with altered risk for Type 2 Diabetes (T2D)^1–5^. Genotype imputation, which predicts genotypes which are not directly measured based on a reference panel, has been widely used for the largest GWAS meta-analyses, but has typically limited imputation to variants with MAF > 0.005. Therefore, large-scale meta-analyses to date have restricted their analysis to variants with MAF larger than 0.005. Until recently, the only approach to analyze variants with MAF below this threshold has been either via whole-genome sequencing (WGS) or whole-exome sequencing (WES). WGS and WES datasets are still limited in sample size, and WES focuses only on the ∼1% of the genome that encodes for protein-coding exons. The National Heart, Lung, and Blood Institute (NHLBI) Trans-Omics for Precision Medicine (TOPMed) imputation reference panel has demonstrated the ability to accurately impute variants with frequency as low as 5×10^−5^ ^5–7^, providing a unique opportunity to explore the contribution of rare and low-frequency variants for the first time at scale.

Approximately 0.4% of all diabetes cases are caused by rare variants with very large effect sizes (odds ratios [OR] often > 20)^8–10^, classified as monogenic diabetes (MD). Many monogenic diabetes subtypes are well suited for precision medicine interventions^11^, but the vast majority of patients with MD are undiagnosed^12^, with populations of non-European genetic ancestry more vulnerable to misdiagnosis^13^. A large percentage of people likely have inherited rare variants with intermediate penetrance, which increases diabetes risk by ∼5-10-fold^14^; however, without clear evidence for the increased risk, clinicians are unable to incorporate genetic considerations in patient care or clinical decision-making. The ability to estimate the pathogenicity of such variants, which are reported as *Variants of Uncertain Significance* (VUS)^15^, is partly based on population-based allele frequency from databases, such as gnomAD^16^, or bioinformatics tools that are used to predict pathogenicity. Segregation in families or enrichment of carriers among those with the disease is used, but this information is often not available or generated in a systematic way.

Estimating the risk of certain variants found in patients of non-European ancestry is particularly limited due to the deficit of genomic data available, such as population-specific allele frequencies^17^. Additionally, it is increasingly recognized that disease risk due to variants of intermediate penetrance can be modified by an individual’s polygenic background, meaning that risk estimation can be improved by combining the rare variants with the polygenic risk scores (PRS) constructed based on common variants^8,14^.

Beyond monogenic diabetes, recent studies using large-scale WGS data have shown that rare variants account for a substantial proportion of the heritability of common complex traits^18^. There is clear evidence for the contribution of rare variants to T2D, and even convergence of signals from rare and common variants^19–21^. The discovery of large-effect, rare, and population-specific variants associated with T2D^3,4,16,17^ suggests a continuum of diabetes subtypes, and degrees of severity rather than a categorical distinction between common forms of diabetes and monogenic diabetes^18^.

We hypothesized that analyzing low-frequency and rare variants at scale would allow for the (1) identification of rare variants of large effect size across diverse ancestries, (2) assessment of the pathogenicity of variants in known MD genes, and (3) investigation of the interplay between rare variants and PRS.

In this study, we combined TOPMed^7^ imputed data from UK Biobank (UKB)^22^, Mass General Brigham Biobank (MGBB)^23^, and Genetic Epidemiology Research on Adult Health and Aging Cohort (GERA)^24^ with WGS data from the All of Us Research Program (AoU)^25^ to perform the largest and most diverse T2D GWAS meta-analysis incorporating variants with MAF as low as 5×10^−5^. The combined dataset comprised 51,256 cases and 370,487 controls, and 12.2% were of non-European ancestry. We further interrogated genetic findings using data from 32 cardiometabolic traits and biomarkers. We identified 8 large-effect variants that are rare or enriched in non-European populations. Finally, we provide a framework for informing pathogenicity based on effect size for diabetes susceptibility with consideration of additional risk stratification using background common genetic variation.

## Results

After quality control and imputation of each of the cohorts with the TOPMed reference panel (except the AoU, as it is WGS data), we conducted a meta-analysis comprising a total of 51,256 cases and 370,487 controls with 12.2% cases of non-European ancestry (Figure 1a, Supplementary Table 1, Supplementary Figure 1). Genotype imputation with TOPMed resulted in a much larger fraction of variants imputed with higher quality, especially at lower allele frequencies, with 10-fold more imputed variants for MAF between 5×10^−5^ and 1×10^−4^ (Supplementary Figure 2) compared to imputation with HRC^26^ and 1000G^27^. We also demonstrate that there is ∼75% minor allele concordance of variants at MAF between 5×10^−5^ and 1×10^−4^ when comparing with data from WES from UKB (Supplementary Figure 3). A comparison of the effect sizes of established variants shows a strong positive correlation (r^2^ ∼ 0.88) in effect sizes between our meta-analysis and published studies (Supplementary Figure 4a). Of note, for low-frequency variants (MAF<0.05), we observed larger effect sizes in our meta-analysis and *p* values of similar significance, despite the effective sample size of our meta-analysis being smaller (180,107 in our study vs 765,591 in Vujkovic et al.^1^), suggesting an improvement of power for low-frequency variants, either because of the more accurate imputation of the TOPMed reference panel or because of more stringent definition of cases and controls (Supplementary Figure 4b,d).

**Figure 1.**
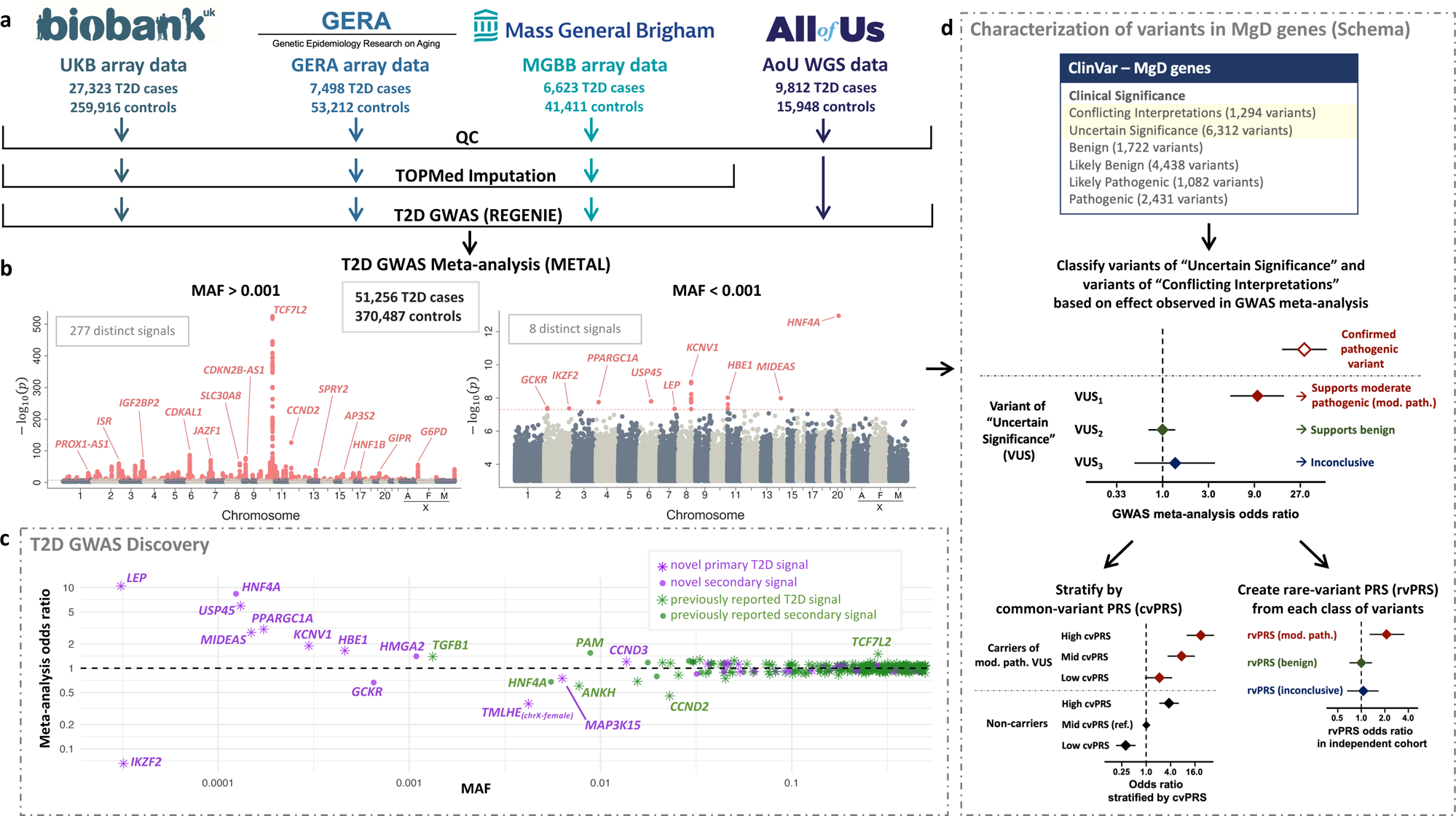
T2D GWAS discovery and overall analysis approach. (a) Overview of the cohorts, sample size, along with the pre-processing steps, for each cohort included in the T2D GWAS meta-analysis. (b) Manhattan plots for variants with an overall study MAF > 0.001 (left) and MAF < 0.001 (right). Pink represents variants that reached genome-wide significance (*p* < 5×10^−8^). (c) Odds ratio for all genome-wide significant conditionally independent variants plotted across MAF. Novel and known signals are represented in purple and green, respectively, and primary and secondary signals as stars or circles. (d) Overview of the downstream analysis that uses rare variant meta-analysis GWAS results to inform the classification of variants in MD genes within several ClinVar categories. We selected all the variants observed in ClinVar in genes involved in monogenic diabetes. For those that are present in our meta-analysis, we categorized variants as “supporting moderate pathogenic” (mod. path), “supporting benign” and “inconclusive” according to the odds ratio and confidence interval. Variants with sufficient carriers within the mod. path category were stratified in tertiles based on their common variant PRS (cvPRS). Finally, we validated the identified mod. path variants in an external dataset (All of Us), comparing the aggregate effect of the carrier status in rare variant PRSs.

### TOPMed imputation, combined with WGS data increases power to detect rare and population-specific variant associations

By performing approximate conditional and joint association analysis (COJO)^28^, we identified 285 distinct signals in 216 loci (Supplementary Tables 2 and 3). Of those, 240 were common (MAF>0.05), 37 were low-frequency (MAF 0.05-0.001), and 8 were rare (MAF<0.001). 51 out of the 284 total signals were novel, of which 14% (34 variants), 24% (9 variants), and 100% (8 variants) were common, low-frequency, and rare, respectively. 37 out of the 51 novel variants were the lead signals, and 14 variants were secondary as they remained significant (*p* < 5×10^−8^) after approximate conditional analysis (Table 1, Figure 1b-c, Supplementary Tables 4 and 5, Supplementary Figure 5).Novel rare (overall discovery MAF <0.001) variants are summarized in Table 1, and Supplementary Figure 5. Variant in 2:27425274-C-T, enriched in African/African American (AA) (rs113171399, MAF_AA_=0.05, OR = 0.66, 95% confidence interval [CI] = 0.57-0.77, *p* = 4.03×10^−8^) is located around *GCKR*, a known T2D-associated region. Approximate conditional analysis showed that it is distinct from the previously reported *GCKR* signal^29^. Variant 2:213085963-G-A (rs1057163508, MAF_AA_=0.001, OR = 0.07, 95% CI = 0.03-0.17, *p* = 4.24×10^−8^) has a strong protective effect and is more enriched in AA ancestry. Variant in 4:23750157-G-A (rs373676665, MAF_MID_=0.03, OR = 3.0, 95% CI = 2.1-4.5, *p* = 1.79×10^−8^) is located near *PPARGC1A*, in a highly muscle-specific enhancer, and it is predicted to disrupt a binding motif recognized by PPARα^30^ a transcription factor that regulates lipid metabolism in various tissues including muscle (Figure 2a-d, Supplementary Figure 6). Variant 6:99432794-G-A, (rs774461980, MAF_ALL_=0.0001, OR = 5.9, 95% CI = 3.2-11.0, *p* = 1.59×10^−8^) located near the *USP45* gene is also associated with lower total circulating bilirubin (*p* = 0.004) and aspartate aminotransferase (*p* = 0.049) in participants without diabetes in UKB (Supplementary Table 4). Variant in 7:128323039-G-A (rs147287548, MAF_AA_=0.002, OR=10.4, 95%CI=4.5-24.2, *p* = 4.53×10^−8^) is an AA specific variant, located in an enhancer active in adipose tissue and adipose tissue-derived mesenchymal stem cells that interacts with the *LEP* gene. This variant disrupts a binding motif for NFATc transcription factors, which are implicated in adipogenesis^31^, Figure 2e-h, Supplementary Figure 7), and is associated with lower apolipoprotein A levels (*p* = 0.003), and lower HDL cholesterol levels in UKB (*p* = 0.002) in participants without diabetes (Supplementary Table 4). Variant 8:110165438-T-C (rs184889639, MAF_AA_=0.02, OR=1.9, 95%CI=1.5-2.3, *p* = 1.06×10^−9^), near *KCNV1*, is enriched in AA ancestry, and is also associated with higher HbA1c and fasting glucose in the latest MAGIC meta-analysis^32^. Variant 14:73781721-C-T (rs190043928, MAF_AA_=0.01, OR = 2.8 95% CI = 2.0-3.9, *p* = 1.03×10^−8^), near *MIDEAS*, is also enriched in AA ancestry and is associated with higher HbA1c in the MAGIC European subset^32^.

**Figure 2.**
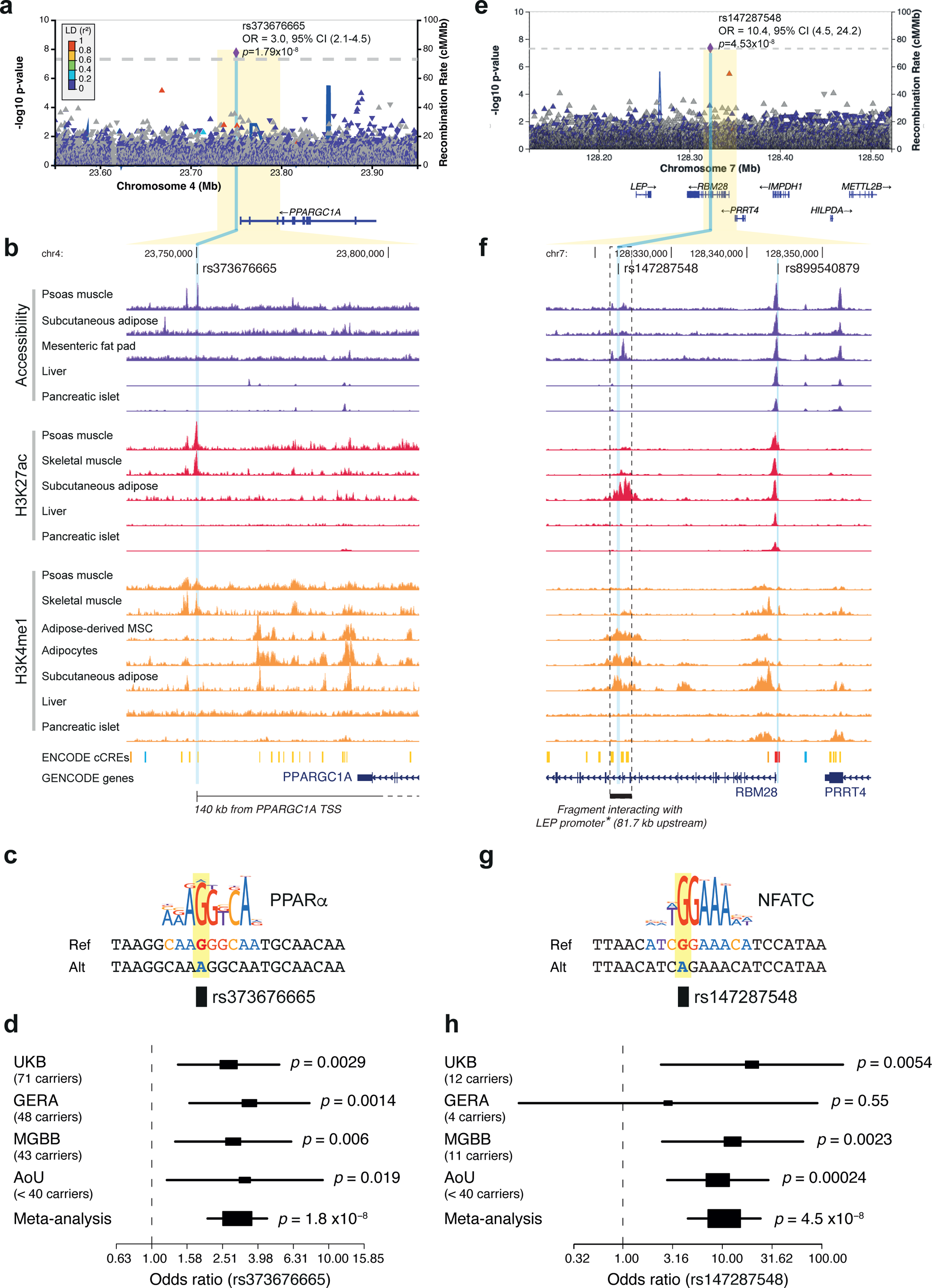
Functional characterization of two novel low-frequency variants associated with T2D. a,b) LocusZoom plots for rs373676665 (a) and rs147287548 (e) regions. Each point represents a variant, with its *p* (on a −log10 scale, y-axis) derived from the meta-analysis association results. The x-axis represents the genomic position (GRC38). (b,f) Representation of chromatin accessibility (ATAC-seq), H3K27ac, and H3K4me1 ChIP-seq signal coverage in T2D-relevant tissues. (b) The box with the dashed line highlights the chromatin fragment that contains rs147287548, which shows significant long-range chromatin interactions with the promoter of the *LEP* gene in mesenchymal stem cells and throughout *in vitro* adipogenesis. The wider chromatin landscape of this locus and chromatin interactions detected by enhancer-capture HiC are shown in Supplementary Figure 7. Details of the datasets shown are provided in Supplementary Table 7. (c) and (g) Transcription factor motif disruption results. The minor alleles of rs373676665 (c) and rs147287548 (g) are predicted to disrupt binding sites for PPARalpha and NFTAc, respectively. (d) Forest plots showing the carrier counts and odds ratios of rs373676665 (d) and rs147287548 (h). Odds ratios are denoted by boxes proportional to the size of the cohort with 95% CI error bars.

**Table 1.** Novel low-frequency variants and/or population-specific variants.

| ID | rsID | EA | NEA | OR | *p* | Direction* | Closest Gene(s) | MAC** | MAF of discovery | MAF gnomAD v3.1.2 |  |  |  |  |
| --- | --- | --- | --- | --- | --- | --- | --- | --- | --- | --- | --- | --- | --- | --- |
|  |  |  |  |  |  |  |  |  |  | AFR | AMR | EAS | EUR | MID |
| 2:27425274 | rs113171399 | T | C | 0.66 (0.57, 0.77) | 4.0E-08 | ---- | *GCKR* | 987 | 0.0007 | 0.046 | 0.004 | 0 | 0.0002 | 0.003 |
| 2:213085963 | rs1057163508 | A | G | 0.07 (0.03, 0.17) | 4.2E-08 | -?-- | *IKZF2* | >12 | 3E-05 | 0.001 | 0 | 0 | 1E-05 | 0 |
| 4:23750157 | rs373676665 | A | G | 3.0 (2.1, 4.5) | 1.8E-08 | ++++ | *PPARGC1A* | >162 | 0.0002 | 7E-05 | 0.0002 | 0 | 0.0004 | 0.035 |
| 6:99432794 | rs774461980 | A | G | 5.9 (3.2, 11.0) | 1.6E-08 | +++? | *USP45* | 73 | 0.0001 | 0 | 0 | 0 | 0.0002 | 0 |
| 7:128323039 | rs147287548 | A | G | 10.4 (4.5, 24.2) | 4.5E-08 | ++++ | *RBM28, LEP* | >27 | 3E-05 | 0.002 | 0.0001 | 0 | 6E-05 | 0 |
| 8:110165438 | rs184889639 | C | T | 1.9 (1.5, 2.3) | 1.1E-09 | ++++ | *KCNV1* | 493 | 0.0003 | 0.023 | 0.005 | 0 | 6E-05 | 0 |
| 14:73781721 | rs190043928 | T | C | 2.8 (2.0, 3.9) | 1.0E-08 | ++++ | *MIDEAS* | 199 | 0.0001 | 0.012 | 0.005 | 0 | 1E-06 | 0.012 |
| 20:44413714 | rs137853336 | T | C | 8.3 (4.7, 14.4) | 1.1E-13 | +++? | *HNF4A* | 71 | 0.0001 | 2E-05 | 0.0002 | 0 | 7E-05 | 0 |

\*Direction of effect in UKBB, GERA, MGB, and AoU, respectively.
\*\*For variants where MAC in AoU is less than 40, MAC is expressed as >N, where N represents the total MAC of UKB+GERA+MGB.

### Association results inform pathogenicity of rare variants in monogenic diabetes genes

Among the genome-wide significant rare variant associations, we highlight a non-synonymous variant (20:44413714-C-T, p.Arg114Trp) in *HNF4A*, a known gene causing Maturity Onset Diabetes of the Young (MODY). The variant p.Arg114Trp is classified as “conflicting interpretations of pathogenicity” in ClinVar (accessed July 2023)^33^, associated with ∼8-fold increased risk of T2D (MAF=0.00007, OR=8.3, 95%CI=4.7-14.14, *p* = 1.08×10^−13^). This variant has previously been reported as a mutation causing a distinct clinical subtype of MD, with reduced penetrance, reduced sensitivity to sulfonylurea treatment, and no effect on birth weight^34^.

Motivated by this example, we analyzed the effect of rare variants in genes known to be involved in MD forms, including MODY, neonatal diabetes, and rare forms of syndromic diabetes (Supplementary Table 6). In a meta-analysis that excluded the AoU cohort, we evaluated the effect of available variants in these genes reported in ClinVar and validated our findings in the AoU cohort. Based on the recommendations by the American College of Medical Genetics (ACMG) guidelines^15^, we defined variants with OR higher than 5 and lower bound (LB) of the 95%CI above 2 as “supporting moderate pathogenic effect”, variants with a 95% CI upper bound (UB) below 2 as “benign,” and variants with 95%CI UB higher than 2 and LB lower than 2 as “inconclusive” as they are still probably too rare to have sufficient power to identify an association if present (Figure 3a).

**Figure 3.**
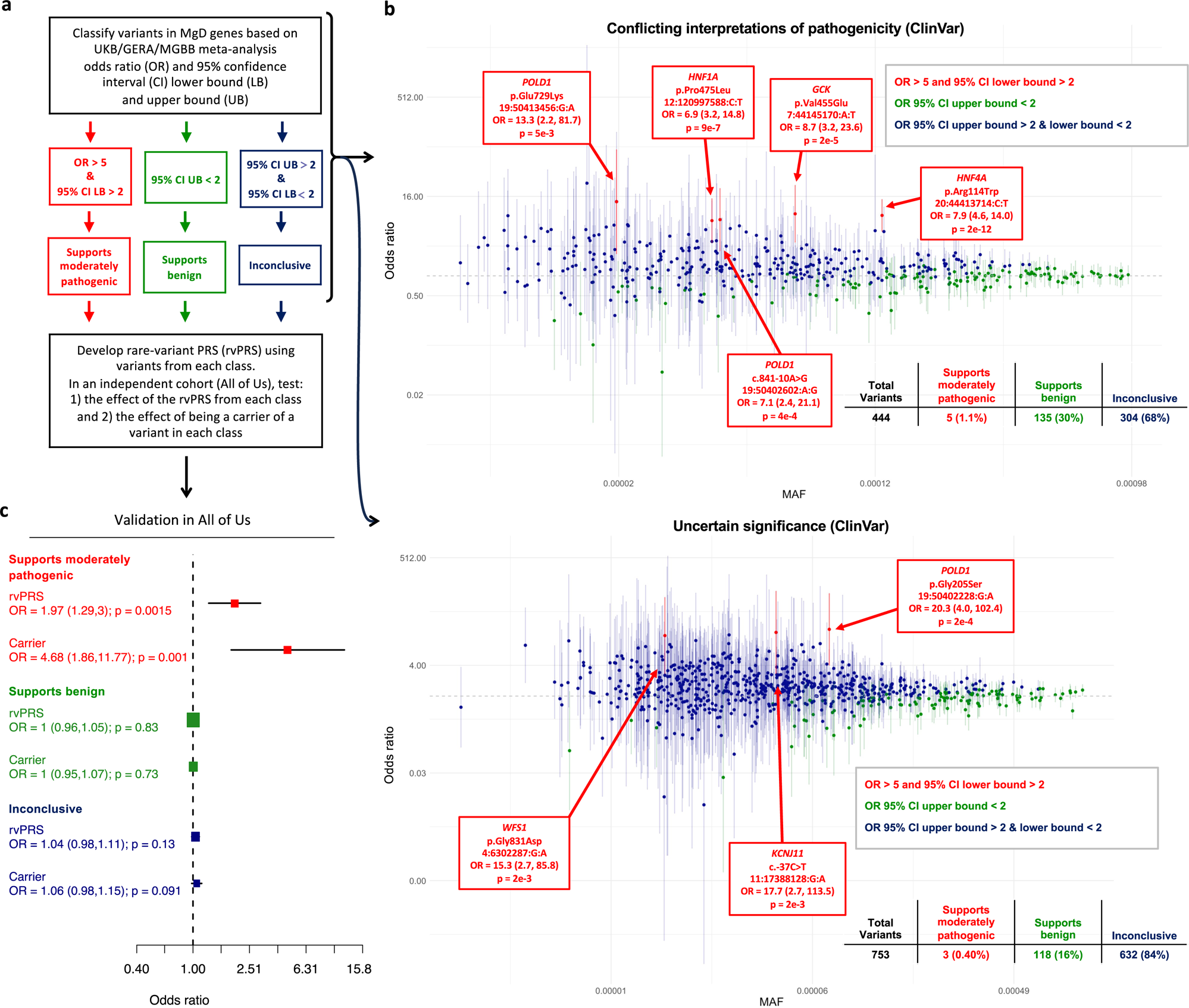
Classification of variants in MD genes from ClinVar and development of rare-variant PRS. (a) Overview of variant classification according to meta-analyses results in UKB/GERA/MGBB (excluding All of Us). We extracted variants in MD genes from ClinVar labeled as “uncertain significance”, “conflicting interpretations of pathogenicity”, “likely benign”, or “likely pathogenic”. We then classified these variants based on the UKB/GERA/MGBB meta-analysis odds ratio (OR) and 95% confidence interval (CI) lower bound (LB) and upper bound (UB). Variants with a meta-analytic OR > 5 and an OR 95% LB > 2 are classified as “supports moderately pathogenic” (red). Variants with an OR 95% UB < 2 are classified as “supports benign” (green). Variants with an OR 95% UB > 2 and LB < 2 are classified as “inconclusive” (blue). (b) Results of this analysis for variants of “conflicting interpretations of pathogenicity” and “uncertain significance” according to ClinVar. The x-axis represents MAF, and odds ratios are represented in the y-axis. Only variants with MAF<0.001 were considered for this analysis. (c) Variants identified as “moderately pathogenic” were aggregated in a rare variant polygenic risk score (rvPRS), a weighted sum of the effect alleles, and tested in AoU. OR for each unit of the rvPRS is represented in addition to the OR associated with being a carrier of any of the risk alleles (Carrier). The forest plots are represented for the group of variants classified as “supports benign”, “supporting moderately pathogenic” and “inconclusive”.

Among the variants within the ClinVar category of “Conflicting interpretation of pathogenicity (CIP)”, we identified 5 variants with OR>5 and 95%CI LB >2 (Figure 3b, Supplementary Figure 7). In addition to the p.Arg114Trp described above, we identified two additional variants located in two known MODY genes, one in *HNF1A*, enriched in Ashkenazi Jewish (AJ) (12:120997588:C:T, p.Pro475Leu, maxMAF_AJ_=0.00086, OR=6.9, 95%CI=3.2-14.8, *p* = 9×10^−7^) and *GCK* (7:44145170:A:T, p.Val455Glu, maxMAF_Eur_=0.00002, OR=8.7, 95%CI=3.2-23.6, *p* = 2×10^−5^). We also identified two variants in the *POLD1* gene (19:50413456:G:A, p.Glu475Lys, maxMAF_Finn_=0.00019, OR=13.3, 95%CI = 2.2-81.7, *p* = 5×10^−3^; 19:50402602:A:G, c.841-10A>G, maxMAF_Eur_=0.00008, OR=7.1, 95%CI=2.4-21.1, *p* = 4×10^−4^). Mutations in *POLD1* have been observed in a multi-system disorder with lipodystrophy causing diabetes^35^. In this same ClinVar group, 135 variants (30% of variants tested) showed ORs with a 95% CI UB below 2, suggesting that these variants could be considered benign despite being currently in the category CIP in ClinVar.

Within the variants the ClinVar category of “uncertain significance” we identified three additional variants (Figure 3b, Supplementary Figure 8), in *WFS1* gene (4:6302287:G:A, p.Gly831Asp, maxMAF_Eur_=0.00002, OR=15.3, 95%CI = 2.7-85.8, *p* = 2×10^−3^), in *KCNJ11* (11:17388128:G:A, 3’UTR, maxMAF_AFR_=0.00002, OR = 17.7, 95%CI = 2.7 - 113.5), *p* = 2×10^−3^) observed only in one carrier in gnomAD in the African/African American subgroup, and in *POLD1* (19:50402228:G:A, Gly205Ser, maxMAF_Eur_=0.00001, OR=20.3, 95%CI=4.0-102.4, *p* = 2×10^−4^). Additionally, 118 (16%) of the variants had association results supportive of benign, based on the lack of association with T2D, informing pathogenicity of these variants.

We identified one additional variant supporting pathogenicity in *POLD1* (19:50409504:C:T, c.2007-15C>T, maxMAF_Eur_=0.00002, OR=9.9, 95%CI=2.1-47.8, *p* = 4×10^−3^) in the ClinVar “likely benign” category. In this category, 258 (33%) were supportive of benign (Supplementary Figure 9). There were no variants with association results supporting pathogenicity in the “likely pathogenic” and “pathogenic” categories (Supplementary Figure 10, 11), probably because such variants have much lower allele frequency and there is still not enough power to detect association in our dataset. However, our results supported evidence of being benign for two variants categorized as “pathogenic” in ClinVar (Supplementary Figure 10). As expected, within the ClinVar “benign” category, 70% of the variants are supported as benign and none supported pathogenicity according to our association data.

To further validate the effects of the variants identified as “supporting moderate pathogenic effect”, we generated a rare variant PRS (rvPRS) using the effect sizes from these variants in a meta-analysis excluding the AoU cohort. Then, we used the AoU cohort as a validation dataset. Each unit of the PRS was associated with two-fold increased risk for T2D (OR=1.97, 95%CI=1.29-3, *p*=0.0015. In addition, we observed that carriers of these variants had 4.7-fold increased risk for T2D (OR=4.68, 95%CI=1.86-11.77, *p*=0.001) versus non-carriers in the AoU cohort. Variants that we classified as “supporting benign” and those that remain inconclusive according to our analysis were not significantly associated with T2D in the AoU cohort (Figure 3c).

### Common variant polygenic risk scores modulate the effect of rare variants in monogenic diabetes genes

To understand potential causes for the *conflicting interpretation of pathogenicity* of the identified variants which support “moderate pathogenic effect” in the three established MODY genes, *HNF4A*, *HNF1A* and *GCK,* we tested how a common variant polygenic risk score (cvPRS) for T2D influences diabetes risk pathogenicity and compared the effects with those of established confirmed MODY variants identified using whole-exome sequence data from the UKB^36^. For the *HNF4A* p.Arg114Trp variant, when compared to non-carriers in the middle tertile of the PRS, carriers in the highest tertile had a higher odds ratio (OR = 18.3, 95%CI = 7.2 - 46.9, *p*=1.2×10^−9^), with an effect size that was comparable to the effect observed in carriers of confirmed pathogenic MODY variants (OR = 17.7, 95%CI= 6.5 - 48.1, *p*=7.8×10^−6^). However, *HNF4A* p.Arg114Trp carriers within the lowest tertile of the PRS showed a much smaller odds ratio (OR=2.62, 95%CI=0.97-7.09, *p*=0.06) (Figure 4a).

Similar to what we observed for the *HNF4A* variant, carriers of either p.Pro475Leu in *HNF1A* and p.Val455Glu in *GCK* who also were in the top tertile of the cvPRS had marked increased risks of diabetes (OR’s > 10). However, carriers in the top tertile had odds ratios smaller than those observed with confirmed *HNF1A* or *GCK* MODY variants (Figure 4b,c).

**Figure 4.**
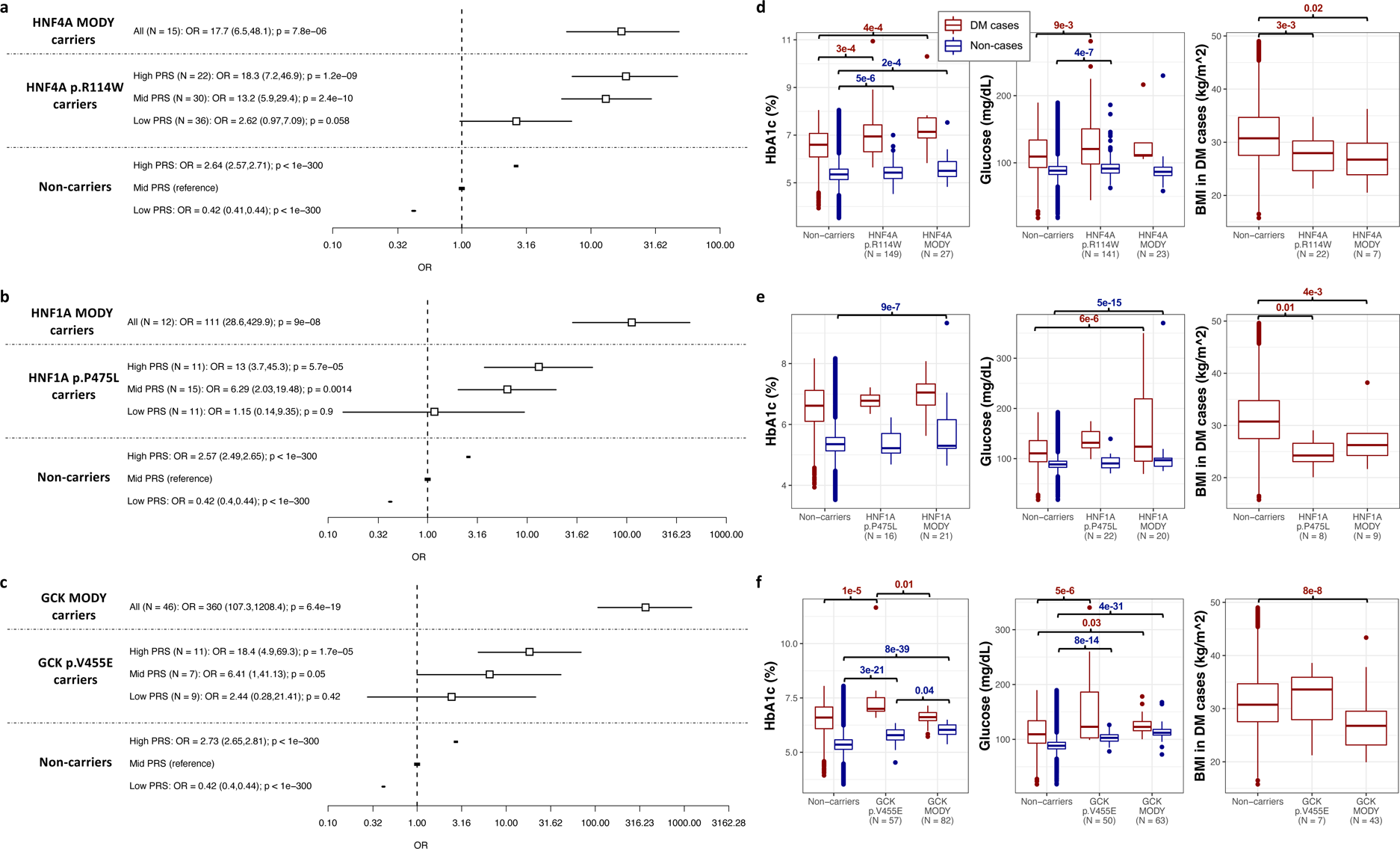
Effect of moderately pathogenic variants versus confirmed pathogenic MODY variants for diabetes risk and on related clinical variables. (a-c) Forest plots showing the effect of p.Arg114Trp, p.Pro475Leu, and p.Val455Glu, stratified by common variant PRS (cvPRS) tertiles. Odds ratios (OR) are denoted by boxes proportional to the size of the cvPRS subgroup and 95% CI error bars. OR are relative to non-carriers in the middle tertile of the cvPRS. On the top of each forest plot, the effect of being a carrier for a confirmed pathogenic variant for HNF4A (a), HNF1A (b), and GCK (c) MODY is also represented, using data identified via exome sequencing in UK Biobank. For each effect estimate, the diabetes case definition included individuals with type 1 or type 2 diabetes. Non-carriers with PRS in the middle tertile were treated as the reference group. (d, e,f) Boxplots of HbA1c (%), random glucose (mg/dL), and BMI (kg/m^2^) in diabetes cases (red) and non-cases (blue) among non-carriers (left), carriers of a moderately pathogenic variant (middle), and carriers of confirmed pathogenic MODY variants (right) in HNF4A (d), HNF1A (e), and GCK (f). The covariate-adjusted *p* is included for comparisons with significant differences (*p* < 0.05) between groups.

### Variants with moderate pathogenic effect show phenotypic characteristics that are intermediate between T2D and MODY

We then tested whether carriers of variants “supporting moderate pathogenic effect” showed a different pattern compared to T2D and MODY. For the three variants, the effect on glycated hemoglobin a 1c (HbA1c) and “random” glucose was in between to those of non-carriers and carriers of confirmed MODY variants, supporting that these may be variants with intermediate penetrance. Carriers of p.Arg114Trp *HNF4A* and p.Pro475Leu *HNF1A* with diabetes had significantly lower body-mass index (Figure 3d,e,f). Carriers of the p.Arg114Trp *HNF4A* variant had significantly lower age at onset of diabetes. These results support that these variants are in a continuum spectrum of pathogenicity in MD.

## Discussion

Genetic studies for T2D have been usually categorized as rare variant association studies (RVAS) or common variant association studies (CVAS)^37^, with RVAS typically performed via WES or WGS, and CVAS typically done via GWAS meta-analyses, since the genotyping array data and imputation panels used to cover only relatively common frequencies. Here, we performed the largest T2D GWAS meta-analysis, including variants with allele frequencies below 0.001, by combining TOPMed imputation from three large and diverse cohorts with WGS data from the *All of Us* Research Program cohort. We demonstrated that TOPMed imputation provides accurate genotypes for variants with MAF as low as 5×10^−5^. We showed that TOPMed imputation is a reliable strategy to analyze low-frequency and rare genetic variation in cohorts for which WGS data is not available. Despite new WGS data at scale becoming available, there is still substantial extant data that can benefit from imputation with large reference panels such as the TOPMed panel, especially from data from non-European ancestries.

This study has enabled several discoveries. First, we identified 55 novel T2D-associated variants, of which 10 were low-frequency (MAF<0.05), and 9 additional were rare or population-specific. For example, the variant rs373676665 is near the *PPARGC1A* gene, which is highly intolerant to loss of function (pLI = 1, gnomAD v2.1.1). *PPARGC1A* codes for peroxisome proliferator-activated receptor gamma coactivator-1 alpha (PGC-1 α), a protein that regulates PPARgamma function^38^, which in turn regulates genes involved in energy metabolism^39^. Integration of T2D-relevant epigenomic maps enabled us to observe that rs373676665 resides in an enhancer that is highly specific to muscle tissue, which is in agreement with previous reports showing a causal link between PGC-1α dysregulation in skeletal muscle, insulin resistance and T2D^40^. Another variant, rs147287548, is located near the *LEP* gene, which encodes for leptin, a hormone/adipokine primarily secreted by white adipose tissue and whose deficiency causes insulin resistance^41^. rs147287548 overlaps with a predicted enhancer active in adipocyte-derived mesenchymal stem cells and adipose tissue that interacts with a region proximal to the *LEP* transcription start site by enhancer-capture HiC^42^, and is predicted to disrupt a NFATc motif. NFATc transcription factors have been implicated in adipogenesis, are modulated in obesity, and regulate glucose and insulin homeostasis^31^. Homozygous mutations in the *LEP* gene are additionally known to cause extreme obesity in human and mouse^43^.

This study also represented a proof of concept of how using large-scale biobank or population-based data can provide additional evidence for pathogenicity of variants within genes known to cause monogenic diabetes. Monogenic diabetes exemplifies precision medicine^44^, as identifying the gene causing diabetes guides the management of the disease. For example, individuals with MODY caused by *HNF1A* mutations can often be transitioned from insulin therapy to oral hypoglycemic agents such as sulfonylureas^45^, and those with MODY caused by *GCK* mutations develop stable mild hyperglycemia and do not need pharmacological therapy as they are often refractory to it^46^ and their risk of microvascular and macrovascular complications is considered low^47^. The current assessment of the pathogenicity of genetic variants relies on bioinformatic prediction, population allele frequency, and expert panel curation. The presence of carriers among patients with diabetes or segregation analyses are also considered, but these data are usually unavailable. For this reason, many of the missense variants identified in these genes remain as variants of uncertain significance or “conflicting interpretation”. By testing rare variant associations at scale using TOPMed imputation, we identified variants in known monogenic diabetes genes with intermediate pathogenicity, which we defined as having an odds ratio higher than 5 and a 95%CI lower bound above 2. Additional evidence may be needed to confirm the pathogenicity and clinical utility of such variants, such as testing whether carriers of *HNF1A* or *HNF4A* variants of intermediate penetrance have heightened response to sulfonylureas. Nevertheless, we propose that our approach could serve as a tool to prioritize variants for future functional validation or deep phenotypic characterization.

Some of the variants we identified as having intermediate penetrance have published evidence of having functional consequences. For example, the *GCK* p.Val455Glu is located in the same position as a GCK activating mutation (p.Val455M) causing familial hyperinsulinism^48^. It has been reported previously in individuals with impaired glucose tolerance or fasting hyperglycemia^49^, and two independent studies have demonstrated that the variant is kinetically inactivating consistent with loss of function^50,51^. Identifying carriers of GCK inactivating mutations is important, not only because these carriers may not need treatment, but also because of possible complications in pregnancy if the risk allele is not inherited by the fetus of carrier mothers, resulting in hyperglycemia and excessive growth in the fetus^52^.

We also demonstrated that carriers of variants identified as “supporting moderate pathogenicity” had a 5-fold increased risk of being diagnosed with T2D in an independent WGS-based cohort, demonstrating that our approach provides robust identification of rare variants with large effect and that rare variants contribute to the overall burden of T2D.

While we were only able to identify a handful of variants that support moderate pathogenicity, we also found a much larger fraction of variants that are sufficiently powered and which, based on their lack of association with diabetes, are supportive of being benign. These results can de-prioritize candidate variants and genes in patients suspected of having monogenic diabetes. An illustrative example is a variant in HNF1A, currently classified as VUS (12-120994274-A-C, p.Glu275Ala, MAF=0.00002, OR=0.65, 95%CI=0.23-1.86, *p*=0.42). This variant is rare, predicted to be deleterious by several bioinformatic tools, and therefore remains as a VUS in ClinVar. However, knowing that carriers of this variant are not at increased risk for T2D can add additional evidence for this variant as being benign.

Our study design also allowed us to assess the interplay between common and rare variants in monogenic diabetes genes. We observed that stratifying carriers of such variants by their cvPRS identifies carriers that can have the same magnitude of increased risk of developing diabetes as the carriers of well-established MODY variants. For example, our results showed that one-third of the carriers of p.Arg114Trp *HNF4A* variant, i. e. those in the highest tertile of the cvPRS, had the same risk as those carrying confirmed MODY variants. In addition, we show that a rare variant PRS (rvPRS) using variants identified as “supporting moderate pathogenic effect” is associated with T2D in an independent cohort (AoU), with carriers of any of such variants showing nearly 5-fold increased risk for T2D. Future work should leverage these data to develop rare variant polygenic risk scores (rvPRS) with the inclusion of variants genome-wide beyond the set of currently known monogenic diabetes genes. However, novel methods that allow modeling linkage disequilibrium between these rare variants and the common variants are needed to fully take advantage of both common and rare genetic variation.

Previous studies have proposed the use of association with endophenotype biomarker data, such as HbA1c, to support pathogenicity^53^. However, association with common forms of diabetes (i. e. T2D) has not been previously used to assess pathogenicity due to the lack of power of WGS or WES based studies.

Our study has several limitations. First, we acknowledge that the standard genome-wide significant threshold (*p* < 5×10^−8^) may not be sufficiently stringent, developed initially as a genome-wide significant threshold for common variants, given the fact that many more variants, including rare and population-specific variants are being tested. However, we believe that there is complementary evidence to support the findings. For example, the variant near the *LEP* gene is located in an enhancer that interacts with the LEP promoter and is also associated with lower apolipoprotein A levels and lower HLD cholesterol, the variant near *KCNV1* is associated with higher HbA1c and fasting glucose in the latest MAGIC meta-analysis^32^, and there are higher priors for missense variants observed in monogenic diabetes genes. In addition, while we have used a broad definition of T2D as a surrogate phenotype to assess potential pathogenicity of monogenic diabetes genes, we cannot discard the possibility that some of the variants have a phenotype that is closer to prediabetes or type 1 diabetes (T1D) rather than T2D. While power is limited to test the association of such rare variants with T1D, our definition of controls excluded those participants who met the criteria for diagnosing T1D or prediabetes. This could only bias the association results towards the null and not increase our false positive rate.

The implementation of these data as an additional metric to assess variant pathogenicity will require additional investigation involving expertise from clinicians, genetic counselors, and the ClinGen Monogenic Diabetes Expert Panel. Nevertheless, we make the full summary statistics available to the scientific community through the Type 2 Diabetes Knowledge Portal (T2DKP) as a resource that can be used for further investigations^54^.

In summary, our work underscores the value of combining WGS data with low-frequency and rare genotype imputation in cohorts for which WGS or WES data still does not exist, not only to discover new associations but also to guide the interpretation of variant pathogenicity. Expanding this work to larger and more diverse populations will contribute to reduce health disparities in the application of precision medicine in diabetes. The framework provided here will serve as an example for the study of genetic variation associated with common and monogenic forms of disease.

## Supplementary Methods

### Description of cohorts and case-control definitions

#### UK Biobank

The UK Biobank (UKB) is a prospective cohort study with genetic and phenotypic data collected on approximately 500,000 individuals across the United Kingdom between 40 and 69 years of age at recruitment ^22^. Participants agreed to provide detailed information about their lifestyle, environment, and medical history, biological samples (for genotyping and biochemical assays), to undergo measures, and to have their health followed (http://www.ukbiobank.ac.uk/). The UKB has obtained ethical approval covering this study from the National Research Ethics Committee (REC reference 11/NW/0382).

Using the UK Biobank array data, we applied the quality control performed by UKB. We filtered the array data to only include variants and samples that were used as input for phasing and present in both arrays after the UKB quality control. This resulted in a total of 687,342 variants and 486,757 samples included for phasing and imputation. We then performed phasing using SHAPEIT4^55^, and imputed the phased haplotypes using the TOPMed reference panel freeze 8^7^. The UKB data was accessed through the application number 27892. UKB is composed of 46% males, 54% females, and ranges in age from 37 to 80 years old with an average age of 57 years. 15% of the individuals are from genetically-inferred non-European ancestry, with 1.4% AFR, 0.20% AMR, 1.8% CSA, 0.55% EAS, 0.3% MID, and 10.8% Other.

Genotype calling was performed by Affymetrix on two closely related purpose-designed arrays. ∼50,000 participants were run on the Applied BiosystemsTM UK BiLEVE AxiomTM Array and the remaining ∼450,000 were run on the Applied BiosystemsTM UK Biobank AxiomTM Array. The dataset combines results from both arrays and there are 805,426 markers in the released genotype data.

We converted the array data to VCF format and performed phasing using SHAPEIT4. We performed phasing separately for each genotyping array (UK BiLEVE versus UK Biobank Axiom array). To ensure appropriate phasing of the X chromosome, we updated the array data to ensure males were encoded as diploid in the VCF file before running SHAPEIT4.

After phasing, we split the data into chunks of about 25,000 samples each, randomly selected independently of ancestry or T2D status. We submitted the chunks to the TOPMed imputation server (https://imputation.biodatacatalyst.nhlbi.nih.gov/)^56^ and, once completed, we merged the imputed data using BCFTOOLS^57^. We filtered the imputed data to exclude variants with an Rsq below 0.3 within each chunk. When merging the chunks after imputation, all variants that were present in any of the imputed chunks were included in the fully merged data. After merging, we converted the VCF files to BGEN format for analysis with REGENIE^58^.

UKB included a total of 27,323 T2D cases and 259,916 controls. We defined T2D cases and controls using an algorithm designed specifically for UKB^59^. Using this algorithm, we defined cases and controls using the following criteria:

a) Case selection criteria.
1) possible T2D
2) probable T2D or
3) had maximum HbA1c above 6.5 and were not possible or probable type 1 diabetes
b) Control selection criteria.
1) not possible or probable type 1 or type 2 diabetes
2) did not have a family history of diabetes
3) had a maximum HbA1c below 5.7 and
4) were older than 45 years of age

#### Mass General Brigham Biobank

Mass General Brigham Biobank (MGBB; formerly Partners HealthCare Biobank)^23^ is a large repository of biospecimens and data linked to extensive electronic health record data and survey data. Its objective is to support and enable translational research focused on genomic, environmental, biomarker, and family history associations with disease phenotypes. MGBB has enrolled more than 135,000 participants and has generated genomic data on more than 65,000 of its participants. MGBB consists of consented patients seen at various U.S. hospitals, including Massachusetts General Hospital, Brigham and Women’s Hospital, McLean Hospital, and Spaulding Rehabilitation Hospital. Patients are recruited in the context of clinical care appointments at more than 40 sites. MGBB participants provide consent for the use of their samples and data in broad-based research. The approval for the analysis of MGB data was obtained from the MGB Institutional Review Board (study 2016P001018). T2D status was defined based on “curated phenotypes” developed by the MGBB Portal team using both structured and unstructured electronic medical record data and clinical, computational, and statistical methods. Natural Language Processing was used to extract data from narrative text. Chart reviews by disease experts helped identify features and variables associated with particular phenotypes and were also used to validate results of the algorithms. The process produced robust phenotype algorithms that were evaluated using metrics such as positive predictive value (PPV) and negative predictive value (NPV)^60^.

We accessed Multi-Ethnic Genotyping Array (MEGA) genotyping data for 36K participants and Infimum Global Screening Array (GSA) genotyping data for 18K participants from MGBB then quality controlled (QCed), phased, and imputed each of these array datasets separately. Briefly, the QC steps included filtering out genotyped variants based on MAF levels (<0.5%), missingness (<0.05), genotyping batch bias (*P*<5×10^−5^), and Hardy-Weinberg equilibrium (*P*<1×10^−10^) and palindromic single nucleotide variants (AT or CG). An additional 2% of individuals were removed if their self-described sex did not match their genetic sex and if they had a high ratio of heterozygote variants. Filtering variants based on Hardy-Weinberg equilibrium and participants based on heterozygosity was performed in self-identified race and ethnicity subgroups. This clean dataset was then phased using Shapeit4 and imputed using the TOPMed reference panel freeze 8. The union of the imputed MEGA and GSA datasets were then merged together into a final dataset.

MGBB included a total of 6,623 T2D cases and 41,411 controls. T2D status was defined based on “curated phenotypes” developed by the MGBB Portal team using both structured and unstructured electronic medical record data and clinical, computational, and statistical methods. Natural Language Processing was used to extract data from narrative text. Chart reviews by disease experts helped identify features and variables associated with particular phenotypes and were also used to validate results of the algorithms. The process produced robust phenotype algorithms that were evaluated using metrics such as positive predictive value (PPV) and negative predictive value (NPV)^60^.

a) Control selection criteria.
1) Individuals determined by the “curated disease” algorithm employed above to have no history of T2D with NPV of 99%.
2) Individuals at least age 55.
3) Individuals with HbA1c less than 5.7
b) Case selection criteria.
1) Individuals determined by the “curated disease” algorithm employed above to have T2D with PPV of 99%
2) Individuals at least age 30 given the higher rate of false positive diagnoses in younger individuals.

Genomic data for 15,061 participants was generated with the Illumina Multi-Ethnic Genotyping Array, which covers more than 1.7 million markers, including content from over 36,000 individuals, and is enriched for exome content with >400,000 markers missense, nonsense, indels, and synonymous variants.

#### Genetic Epidemiology Research on Adult Health and Aging

Genetic Epidemiology Research on Adult Health and Aging (GERA) cohort data was obtained through dbGaP under accession phs000674.v1.p1. For further information about the specific phenotypes (ICD-9-CM codes) included in GERA, visit its website on dbGaP (https://www.ncbi.nlm.nih.gov/projects/gap/cgi-bin/GetPdf.cgi?id=phd004308).

The GERA Cohort was created by a RC2 Grand Opportunity grant that was awarded to the Kaiser Permanente Research Program on Genes, Environment, and Health (RPGEH) and the UCSF Institute for Human Genetics (AG036607; Schaefer/Risch, PIs). The RC2 project enabled genome-wide SNP genotyping (GWAS) to be conducted on a cohort of over 100DK adults who were members of the Kaiser Permanente Medical Care Plan, Northern California Region (KPNC), and participating in its RPGEH. The resulting GERA cohort is composed of 42% males, 58% females, and ranges in age from 18 to over 100 years old with an average age of 63 years at the time of the RPGEH survey (2007).

Briefly, the QC steps included filtering out genotyped variants based on MAF levels (<0.5%), missingness (<0.05), genotyping batch bias (*P*<5×10^−5^), and Hardy-Weinberg equilibrium (*P*<1×10^−10^) and palindromic single nucleotide variants (AT or CG). An additional 2% of individuals were removed if their self-described sex did not match their genetic sex and if they had a high ratio of heterozygote variants. Filtering variants based on Hardy-Weinberg equilibrium and participants based on heterozygosity was performed in self-identified race and ethnicity subgroups.

This study included 60,710 individuals of European ancestry from the GERA cohort. We applied pre-imputation quality control protocol using PLINK^61^. Similar to MGBB, the QC steps for the GERA array data included filtering out variants based on MAF (<0.5%), missingness (<0.05), Hardy-Weinberg equilibrium (*P*<1×10^−10^), and palindromic single nucleotide variants (AT or CG). No individuals were removed due to discordance between self-described and genetically-inferred sex. A total of 647,106 variants and 60,710 samples were included following quality control. For phasing and imputation, we performed the same steps as described above for UKB.

GERA included a total of 7,498 T2D cases and 53,212 controls.

Inclusion criteria:

a) Eligible for RPGEH survey
a) ≥ 18 years of age at time of survey mailing (2007).
b) KP Northern California Region enrollee for at least 2 years prior to survey.
b) Consented to contribute biospecimen to RPGEH and returned saliva sample by cut-off date for GERA genotyping.
c) Successfully genotyped (DQC ≥ 0.82; call rate ≥ 0.97) from extracted DNA.
d) Consented explicitly to have data deposited in NIH-maintained database.

Exclusion criteria:

a. Subject requested withdrawal from study after DNA extraction and genotyping.
b. Validity of link between biospecimen and study participant questionable because of genotype-phenotype discordance, e.g. gender.

A participant was coded as a T2D patient if he/she had at least two diagnoses within this disease category that had to be recorded on separate days. Diagnoses were obtained from patient encounters at Kaiser Permanente Northern California facilities from January 1, 1995 to March 15, 2013.

The March 2013 ICD9-CM diagnoses used for the T2D category were:

a. 250.00 Diabetes mellitus without mention of complication, type II or unspecified type, not stated as uncontrolled.
b. 250.02 Diabetes mellitus without mention of complication, type II or unspecified type, uncontrolled.
c. 250.10 Diabetes with ketoacidosis, type II or unspecified type, not stated as uncontrolled.
d. 250.12 Diabetes with ketoacidosis, type II or unspecified type, uncontrolled.
e. 250.20 Diabetes with hyperosmolarity, type II or unspecified type, not stated as uncontrolled.
f. 250.22 Diabetes with hyperosmolarity, type II or unspecified type, uncontrolled.
g. 250.30 Diabetes with other coma, type II or unspecified type, not stated as uncontrolled.
h. 250.32 Diabetes with other coma, type II or unspecified type, uncontrolled.
i. 250.40 Diabetes with renal manifestations, type II or unspecified type, not stated as uncontrolled.
j. 250.42 Diabetes with renal manifestations, type II or unspecified type, uncontrolled.
k. 250.50 Diabetes with ophthalmic manifestations, type II or unspecified type, not stated as uncontrolled.
l. 250.52 Diabetes with ophthalmic manifestations, type II or unspecified type, uncontrolled.
m. 250.60 Diabetes with neurological manifestations, type II or unspecified type, not stated as uncontrolled.
n. 250.62 Diabetes with neurological manifestations, type II or unspecified type, uncontrolled.
o. 250.70 Diabetes with peripheral circulatory disorders, type II or unspecified type, not stated as uncontrolled.
p. 250.72 Diabetes with peripheral circulatory disorders, type II or unspecified type, uncontrolled.
q. 250.80 Diabetes with other specified manifestations, type II or unspecified type, not stated as uncontrolled.
r. 250.82 Diabetes with other specified manifestations, type II or unspecified type, uncontrolled.
s. 250.90 Diabetes with unspecified complication, type II or unspecified type, not stated as uncontrolled.
t. 250.92 Diabetes with unspecified complication, type II or unspecified type, uncontrolled.

The rest of subjects not coded as T2D patients were considered as controls.

### All of Us research program data

The All of Us Research Program (AoU) is a large, US funded biobank developed to leverage the diversity of the US for facilitating and improving high powered genetic and epidemiological studies^25^. We used the AoU short-read whole genome sequencing (srWGS) data from June 22, 2022 for 98,590 participants (release v6).

AoU provides a list of flagged individuals with poor srWGS data quality based on the number of variants, insertion/deletion ratios, number of insertions, number of deletions, heterozygosity of variants or indels, transition/transversion ratios, and number of variants not in gnomAD v3.1. Additionally, variants from srWGS were flagged as being low quality based on a GQ threshold of <20, a DP threshold of <10, an allele balance filter of <0.2, a low QUAL score (60 for SNPs and 69 for indels), or an ExcessHet score <54.69. These individuals and variants were removed from all subsequent analyses.

Type 2 diabetes cases and controls were defined using a modified version of the Northwestern T2D algorithm ^62,63^. There are 6 conditions for a participant to be determined as having T2D. 1) A participant has no ICD-based diagnosis of T1D AND has an ICD-based T2D diagnosis AND no insulin prescription in an out-patient setting AND is prescribed non-insulin diabetes medications. 2) A participant has no ICD-based diagnosis of T1D AND has an ICD-based T2D diagnosis AND has an insulin prescription in an out-patient setting AND has a non-insulin diabetes prescription prescribed before the insulin. 3) A participant has no ICD-based diagnosis of T1D AND has two separate ICD-based T2D diagnoses AND has an insulin prescription in an out-patient setting AND does not have a prescription for a non-insulin diabetes medication. 4) A participant has no ICD-based diagnosis of T1D AND has an ICD-based T2D diagnosis AND no insulin prescription in an out-patient setting AND is not prescribed a non-insulin diabetes medication AND has an abnormal glycemic lab. 5) A participant has no ICD-based diagnosis of T1D AND has no ICD-based T2D diagnosis AND has a prescription for a non-insulin diabetes medication AND has an abnormal glycemic lab. 6) A participant self-reports as having T2D in their intake survey. Abnormal glycemic labs for cases were defined as a fasting glucose >= 126 mg/dl or a hemoglobin A1c >= 6.5%.

Controls individuals without diabetes were defined if they had 1) at least one in-person healthcare provider visit, 2) at least one glucose measurement available, 3) no abnormal glycemic lab values, 4) no ICD codes or survey responses for any type of diabetes or related conditions, 5) no prescriptions of diabetes medications. We further excluded controls younger than 45.

In total, we identified 9,812 T2D cases and 15,948 controls with complete genetic data and covariate data (age, sex, and BMI and principal components), which we used in our discovery analysis.

#### Assessment of imputation accuracy

We compared the data imputed with the TOPMed reference panel to that of a previous imputation of UKB, which was based on HRC, 1000G, and UK10K. We compared the average INFO score provided by REGENIE, and total number of variants in the TOPMed imputation versus the HRC-1000G-UK10K imputation across the allele frequency spectrum before and after filtering by an INFO score of 0.7 (Supplementary Figure 2). In addition, in a subset of 40K UKB samples with WES data, we evaluated the average percentage of carriers identified in the WES that are identified with the TOPMed imputation across the allele frequency spectrum.

#### Association analysis

We performed whole-genome regression analysis using REGENIE version 2 for UKB, GERA, and MGBB and version 3 for AoU. We used T2D as a binary outcome and included age, sex, body mass index (BMI) and 10 PCs and imputation batch for each of the cohorts. We used a block size of 1,000 for step 1 and a block size of 400 for step 2. All variants with a MAC < 3 for UKB, GERA, and MGBB and < 5 for AoU among T2D cases and controls were excluded from the analysis. To provide better calibrated test statistics, REGENIE supports the option (--firth -- approx --firth-se) to use the Firth correction for variants where the p-value from the standard logistic regression test is below a threshold, as defined by pthresh. For UKB, GERA, and MGBB, we set pThresh to 0.999, causing the Firth correction to be applied for all variants. For AoU, we used the default pThresh of 0.05. After generating summary statistics using REGENIE for each cohort, we used the software METAL^64^ to meta-analyze the results, weighting cohorts by the inverse of the standard error for each variant. Our threshold for genome-wide significance was p < 5 × 10−8.

#### Post-association analysis

##### Identification of the independent set of T2D-associated markers

We used the conditional regression-GCTA (cojo-GCTA)^65^ method on the GWAS meta-analysis summary data. This approach allowed us to assess each locus with a joint combination of several independent markers, corrected for LD between the markers. We constructed an external reference sample with individual genotypes from the UKB and MGBB cohorts to obtain an approximate LD structure. Our reference sample comprised 45,918 samples, ensuring that the proportions of genetic ancestry diversity matched those included in the meta-analysis (88% EUR, 2% AFR, <1% EAS, 1% AMR, 2% SAS). Before the analysis, we filtered the genotypes for imputation quality, retaining those with r2 ≥ 0.8, a minimum dosage certainty of ≥ 0.8 and a MAF ≥ 0.0001%. After filtering, we kept a subset of 78,867,233 markers for performing the conditional analyses. We assumed that markers on different chromosomes or those located more than 2Mb distant from each other were uncorrelated.

#### Criteria for definition of novel variants

To define a novel variant, we followed a two-step procedure. We first looked for each independent signal in the MVP^1^ and the DIAMANTE^2^ multi-ancestry T2D GWAS meta-analyses. Second, for those index variants that were absent or that had an association (*p* < 5×10^−8^), in the Vujkovic et al, in the Mahajan et al 2018, in the Mahajan et al 2022 or in Kurki et al 2023 T2D GWAS meta-analyses, we used our reference sample to extract all tag variants in LD r^2^>0.8 within 2Mb of the index variant. We performed lookups of the tag in any of the T2D GWAS meta-analyses listed above. We defined a novel variant when: i) neither the index nor their tag variants were significantly associated (*p* < 5×10^−8^) in any of the T2D GWAS meta-analyses listed above, or ii) the index variant was not associated in any of the T2D GWAS meta-analyses listed above AND no tag variant was identified.

#### Analysis of variants in monogenic diabetes genes

While more than 40 genes are known to cause MD, distinguishing pathogenic variants from those that are benign in these genes still remains a challenge. Databases such as ClinVar provide designations of variant pathogenicity as determined by submitting laboratories and researchers. However, misclassification of variant pathogenicity is not uncommon, for example in entries that predated the current gold-standard curation approach, as well as for variants that are enriched in understudied populations. The ClinVar designations of variant pathogenicity include: benign, likely benign, likely pathogenic, pathogenic, uncertain significance, or conflicting interpretations of pathogenicity. We evaluated the results in our meta-analysis for variants in ClinVar that are located in known MD genes and have a MAF < 0.001, with a particular focus on variants of uncertain significance (VUSs) and variants with conflicting interpretations of pathogenicity (CIPs). We classify these variants based on the effect observed in the meta-analysis of UKB, GERA, and MGBB, with AoU held out to serve as a validation cohort. We classify the variants based on the meta-analytic odds ratio (OR) and 95% confidence interval (CI) lower bound (LB) and upper bound (UB) (Figure 3). Variants with an OR > 5 and 95% CI LB > 2 were classified as “supports moderately pathogenic”, variants with a 95% CI UB < 2 were classified as “supports benign”, and variants with a 95% CI UB > 2 and 95% CI LB < 2 were classified as “inconclusive”.

#### Polygenic risk scores to stratify carriers of monogenic diabetes variants

We generated a T2D PRS using the PRS-CS software with the auto option used to set the global shrinkage parameter. As input to the software, we meta-analyzed summary statistics from the European ancestry subset GWAS meta-analysis of the T2D published by Vujkovic et al^1^. and the T2D GWAS made publicly available by the FINNGEN Consortium^66^ (Release 6), which did not have overlap with any of the samples included in our meta-analysis. The summary statistics had a final sample size of 277,802 T2D cases and 1,434,249 controls. The meta-analysis was performed using METAL, with inverse-variance weighting. As a linkage disequilibrium (LD) reference panel for PRS-CS we use the LD panel from the UK Biobank, available from the PRS-CS github page. After applying the PRS, we tested the PRS-stratified effect of three rare coding variants, 20:44413714:C:T, 12:120997588:C:T, and 7:44145170:A:T, (Figure 3), that are considered CIPs from the ClinVar designation, but were classified as “supports moderately pathogenic” from the analysis of variants in MD genes described above. Individuals were stratified based on PRS tertiles, with individuals in the top tertile considered to have a “High PRS” and individuals in the bottom tertile considered to have a “Low PRS”. For each effect estimate, the diabetes case definition included individuals with type 1 or type 2 diabetes and individuals who do not carry a variant and have a PRS in the middle tertile were treated as the reference group. This analysis was done separately in each cohort and meta-analyzed with inverse-variance weighting. We also compared the PRS-stratified effects of these three variants with the effect of being a carrier for a confirmed pathogenic variant for *HNF4A*, *HNF1A*, and *GCK* MODY using data from UKB exome sequencing (Figure 3).

#### Testing a rare variant burden in an independent sample in All of Us

Using the variants from each of the three classes described above (“supports moderately pathogenic”, “supports benign”, and “inconclusive”), we developed three rare-variant polygenic risk scores (rvPRSs) and tested the rvPRSs in an independent cohort, AoU (Figure 3). We applied the rvPRS using the --score function in PLINK with the weight for each variant defined as the beta value from the UKB/GERA/MGBB meta-analysis. We evaluate the OR per standard deviation of the score, along with the OR of being a carrier for any of the variants in the score.

#### Biomarker analysis in UK Biobank

Using REGENIE version 2, we performed whole-genome regression of 30 biomarkers in UKB. To ensure normality, we applied rank inverse normal transformation to each of the biomarkers. As the motivation for this analysis was to provide supporting evidence for novel variants identified in the T2D GWAS, we excluded individuals who were identified as T2D cases for the GWAS in UKB. Therefore, the biomarker analysis included the 175,039 individuals in UKB who were identified as controls, along with the 259,916 individuals who were not identified as a T2D case or a control. The 30 biomarkers included: albumin, alkaline phosphatase, alanine aminotransferase, apolipoprotein A, apolipoprotein B, aspartate aminotransferase, direct bilirubin, urea, calcium, cholesterol, creatinine, c reactive protein, cystatin c, gamma glutamyl transferase, glucose, glycated hemoglobin, HDL cholesterol, IGF 1, LDL direct, lipoprotein a, oestradiol, phosphate, rheumatoid factor, SHBG, total bilirubin, testosterone, total protein, triglycerides, urate, vitamin D.

#### Functional in silico interpretation of novel non-coding variants

Lead variants from the GWAS were investigated in combination with all other variants in high linkage disequilibrium (All populations, r2 > 0.8). This expanded set of variants was intersected with ENCODE’s collection of candidate CREs (cCREs) (GRCh38, SCREEN Registry V3), which represented 1,063,878 cCREs across 1,518 cell types. To ascertain tissue and/or cell type-specific of cCREs hosting T2D variants, we used SCREEN (https://screen.wenglab.org/) and the Roadmap Epigenomics 127-reference chromatin states (12-mark, 25-state imputation based, lift-over to GRCh38), which were visualized using the WashU Epigenome Browser^67^. This analysis was complemented with interrogation of human pancreatic islet epigenomic datasets from Miguel-Escalada et al. 2019^68^, which were re-aligned to hg38 assembly for this analysis, and interrogation of epigenomic datasets from human mesenchymal stem cells undergoing adipocyte differentiation from Madsen et al.^42^ and Rauch et al. ^69^, which had coordinates lifted from hg19 to hg38 for the main figure (Figure 2f), and are shown for hg19 in the Supplementary Figure 7. A list of the representative epigenomic datasets from diabetes-relevant tissues used generate Figures 2b,f and Supplementary Figures 6 and 7 using the UCSC Genome Browser^70^ is provided in Supplementary Table 7.

The R package motifbreakR (version: 2.13.7)^71^ was used to carry out motif disruption analysis of variants residing in CREs of diabetes-relevant tissues. The variant data were downloaded from the dbSNP155 database (version: GRCh38_0.99.23) in the human genome (version: hg38_1.4.5) using the snps.from.rsid() command. Transcription factor binding motif data (ENCODE, HOMER, Hocomoco, and FactorBook) were downloaded from the MotifDB database (version: 1.42.0)^72^. The default, weighted sum method was used for disruption analysis where the difference of the probabilities for the two letters of the variant were calculated. The databases were queried for disruptive effect of the variant with p-value maximum 0.001. Results were plotted in R (version: 4.3.1). From the obtained disrupted motifs, we retained those with the flag for ‘strong’ effect and corresponding to transcription factors expressed in the tissues of interest (gene expression queried using the TIGER Data Portal, https://tiger.bsc.es/).

*Functional validation of “novel variants in monogenic diabetes genes” [Anna Gloyn]*

## DATA AVAILABILITY

Full summary statistics of the T2D case-control meta-analysis and the biomarker analysis will be made available through the Common Metabolic Diseases Knowledge Portal (https://cmdkp.org/), and the GWAS catalog (https://www.ebi.ac.uk/gwas/).

## Supplementary Figures

**Supplementary Figure 1.** Comparison of UKB data imputed with TOPMed versus HRC-1000G-UK10K (original imputation release). The line graph shows the average INFO score, and the bar plots show the total number of variants in the TOPMed imputation (blue) versus the HRC-1000G-UK10K imputation (grey) across the allele frequency spectrum before (a) and after (b) filtering for variants with an INFO score greater than 0.7.

**Supplementary Figure 2.** Benchmark of TOPMed imputation accuracy across the allele frequency spectrum. Average percentage of carriers of variants identified in MD WES in Goodrich et al that are identified with imputation in a subset of 40K UKB samples. The y-axis represents the average proportion of carriers identified among variants with imputation INFO > 0.8 in the imputed data from TOPMed vs HRC-1000G-UK10K (original UKB imputation release). The x-axis represents the different allele frequency bins.

**Supplementary Figure 3.** Comparison of UKB/GERA/MGBB/AoU results for lead variants from largest T2D GWAS (Vujkovic et al.) meta-analysis. Comparison of effect estimates (a and b) and −log10(p) values (c and d) from Vujkovic et al. (x-axis) and UKB/GERA/MGBB/AoU meta-analysis (y-axis) for lead variants from Vujkovic et al. Panels a and b compare beta and standard error values for variants with MAF > 0.05 and MAF < 0.05, respectively. The standard error from the UKB/GERA/MGBB/AoU results are represented by the blue vertical bars, while the standard error from the Vujkovic et al. results are represented by the black horizontal bars. Panels c and d compare the −log10(*p*) values for variants with MAF > 0.05 and MAF < 0.05, respectively.

**Supplementary Figure 4.** LocusZoom plots of novel genome-wide significant variants (p < 5 × 10^−8^) and corresponding forest plots showing the carrier counts and odds ratios for each of the meta-analysis cohorts in which the variant was present. Odds ratios are denoted by boxes proportional to the size of the cohort and 95% CI error bars.

**Supplementary Figure 5.** Forest plots showing the carrier counts and odds ratios of each moderately pathogenic variant (odds ratio > 5, and lower-bound 95% > 2) identified in the analysis of variants from ClinVar in MD genes. Odds ratios are denoted by boxes proportional to the size of the cohort and 95% CI error bars.

**Supplementary Figure 6.** Epigenomic landscape of the *PPARGC1A* locus in 127 human tissues and cell types. Colored tracks show Roadmap Epigenomics 12-mark, 25-state imputation-based chromatin state models (GRCh38 lift-over version). The entire topologically associating domain (TAD) containing the rs373676665 association signal (see Figure 2a,b) is shown with the major isoform for each gene represented. The TSS of *PPARGC1A* is located 140 Kb away from rs373676665; while the two other closest transcripts *MIR573* and *DHX15* are located 770 and 834 Kb away, respectively. The zoomed inset at the bottom highlights the only tissues (out of 127) in which the region where rs373676665 resides is annotated as an enhancer.

**Supplementary Figure 7.** Epigenomic landscape of the *LEP* locus. (a) Colored tracks show Roadmap Epigenomics 12-mark, 25-state imputation-based chromatin state models (GRCh38 lift-over version) for 127 human tissues and cell types. The zoomed inset at the bottom highlights the only tissues (out of 127) in which the region where rs147287548 resides is annotated as an enhancer. (b) Chromatin landscape of the LEP locus throughout *in vitro* adipogenesis^1^. The left panel shows all enhancer-capture HiC^1^ chromatin interactions stemming from the fragment containing the rs147287548 variant, which resides in an active enhancer in mesenchymal stem cells and throughout adipogenesis (see also Figure 2f, and panel a of this figure). The right panel shows a zoomed-in region, revealing more clearly chromatin interactions between the rs147287548-enhancer and the promoter of the *LEP* gene.

**Supplementary Figure 8.** Boxplots of HbA1c (%), glucose (mg/dL), BMI (kg/m^2^), and age of diabetes diagnosis in diabetes cases (red) and non-cases (blue) among non-carriers (left) versus carriers of any of the identified moderately pathogenic variants (right) in POLD1.

**Supplementary Figure 9.** Classification of variants in MD genes labeled as “likely benign” in ClinVar. The x-axis represents MAF, and odds ratios are represented in the y-axis. Only variants with MAF<0.001 were considered for this analysis. Variants with a meta-analytic OR > 5 and an OR 95% LB > 2 are classified as “supports moderately pathogenic” (red). Variants with an OR 95% UB < 2 are classified as “supports benign” (green). Variants with an OR 95% UB > 2 and LB < 2 are classified as “inconclusive” (blue).

**Supplementary Figure 10.** Classification of variants in MD genes labeled as “pathogenic” in ClinVar. The x-axis represents MAF, and odds ratios are represented in the y-axis. Only variants with MAF<0.001 were considered for this analysis. Variants with a meta-analytic OR > 5 and an OR 95% LB > 2 are classified as “supports moderately pathogenic” (red). Variants with an OR 95% UB < 2 are classified as “supports benign” (green). Variants with an OR 95% UB > 2 and LB < 2 are classified as “inconclusive” (blue).

**Supplementary Figure 11.** Classification of variants in MD genes labeled as “likely pathogenic” in ClinVar. The x-axis represents MAF, and odds ratios are represented in the y-axis. Only variants with MAF<0.001 were considered for this analysis. Variants with a meta-analytic OR > 5 and an OR 95% LB > 2 are classified as “supports moderately pathogenic” (red). Variants with an OR 95% UB < 2 are classified as “supports benign” (green). Variants with an OR 95% UB > 2 and LB < 2 are classified as “inconclusive” (blue).

