## Supplementary Figures for "Rare variant association analysis in 51,256 type 2 diabetes cases and 370,487 controls informs the spectrum of pathogenicity of monogenic diabetes genes"

Supplementary Figure 1

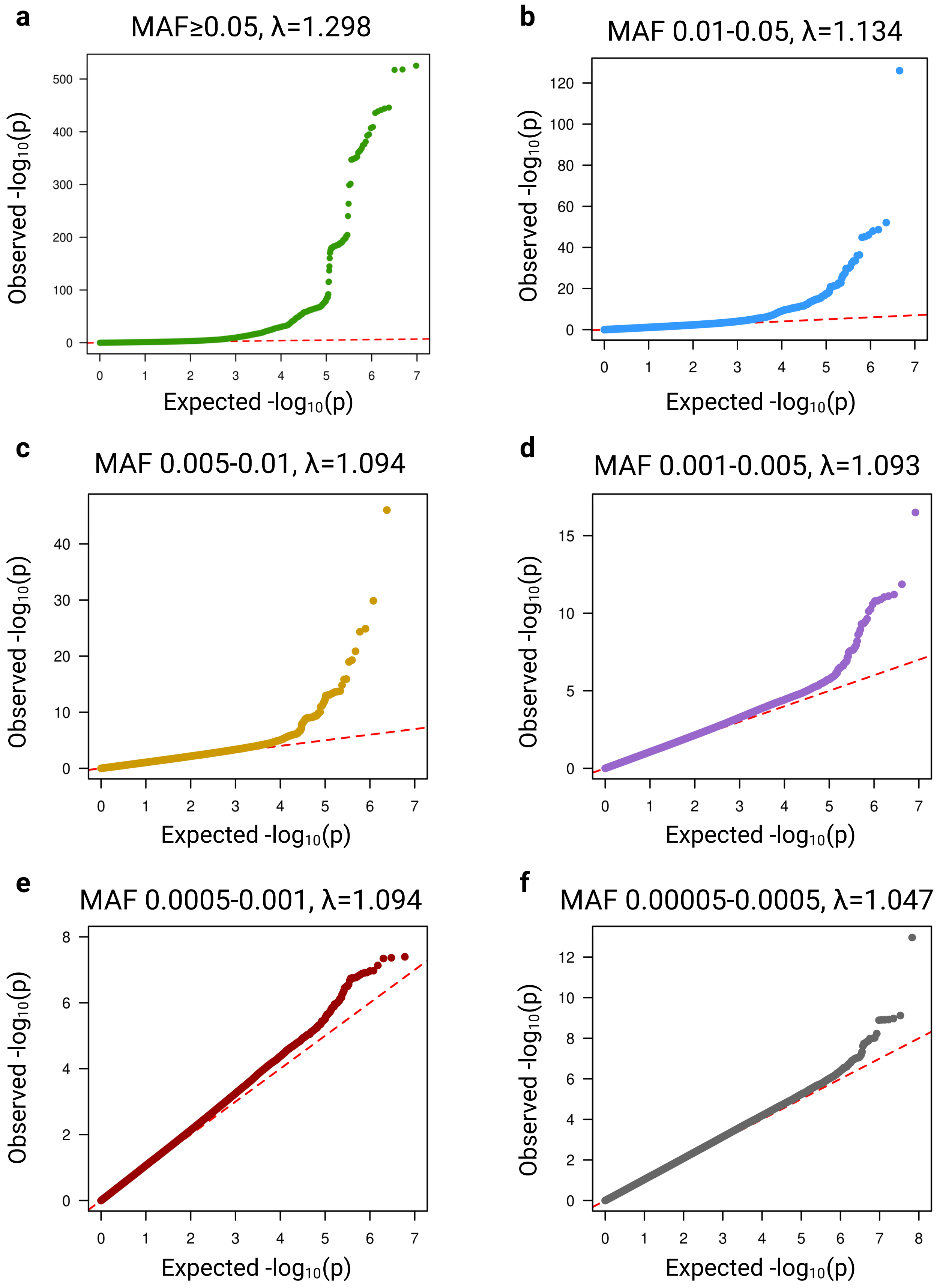

Supplementary Figure 2

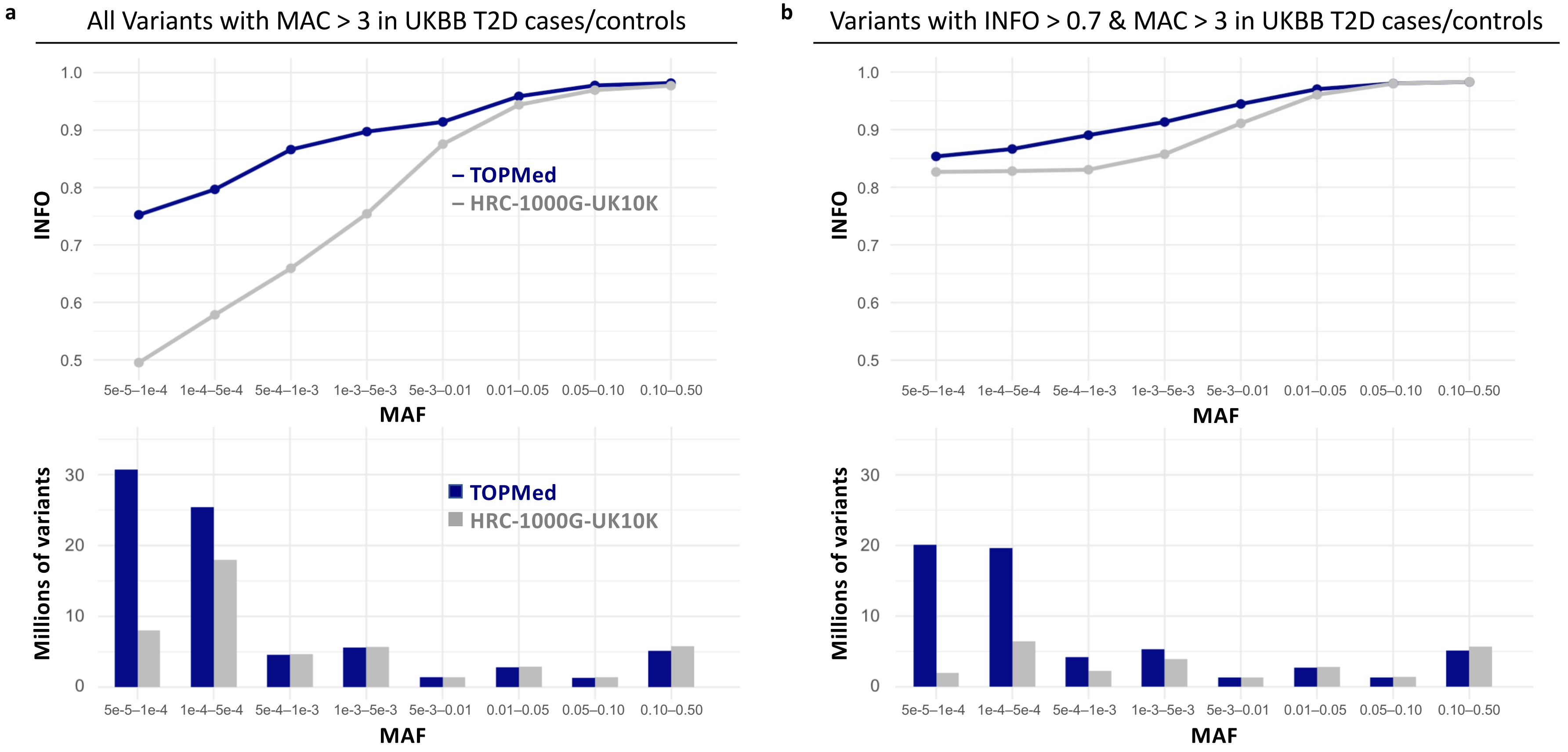

Supplementary Figure 3

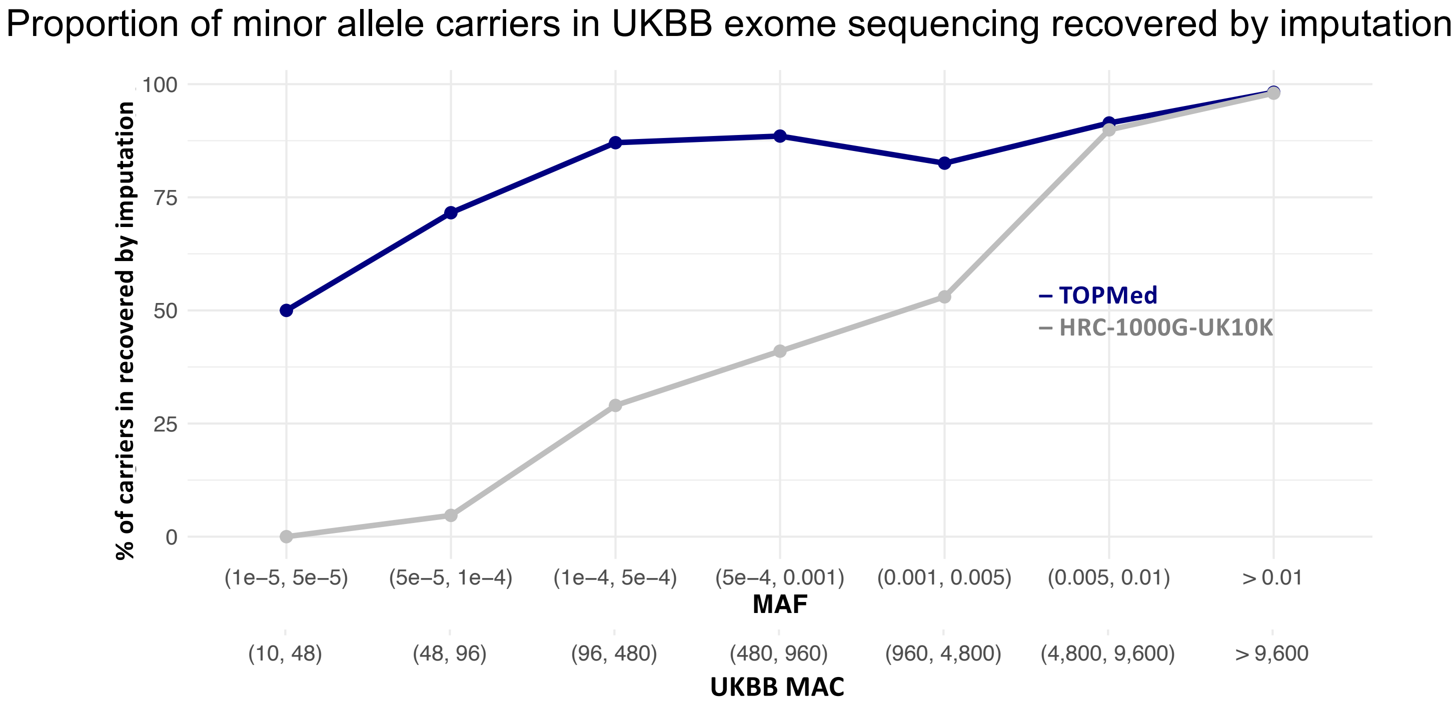

### Supplementary Figure 4

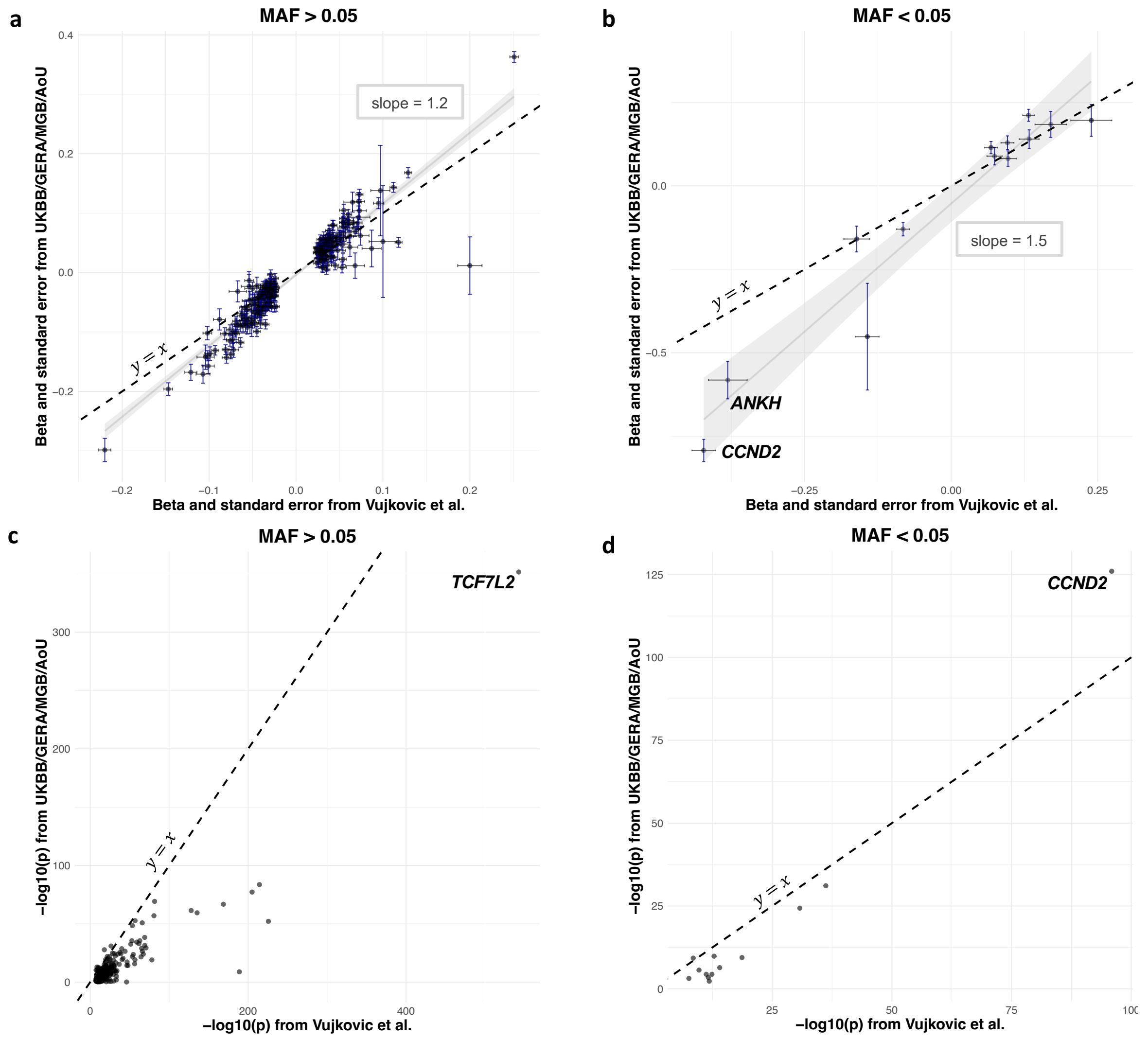

### Supplementary Figure 5

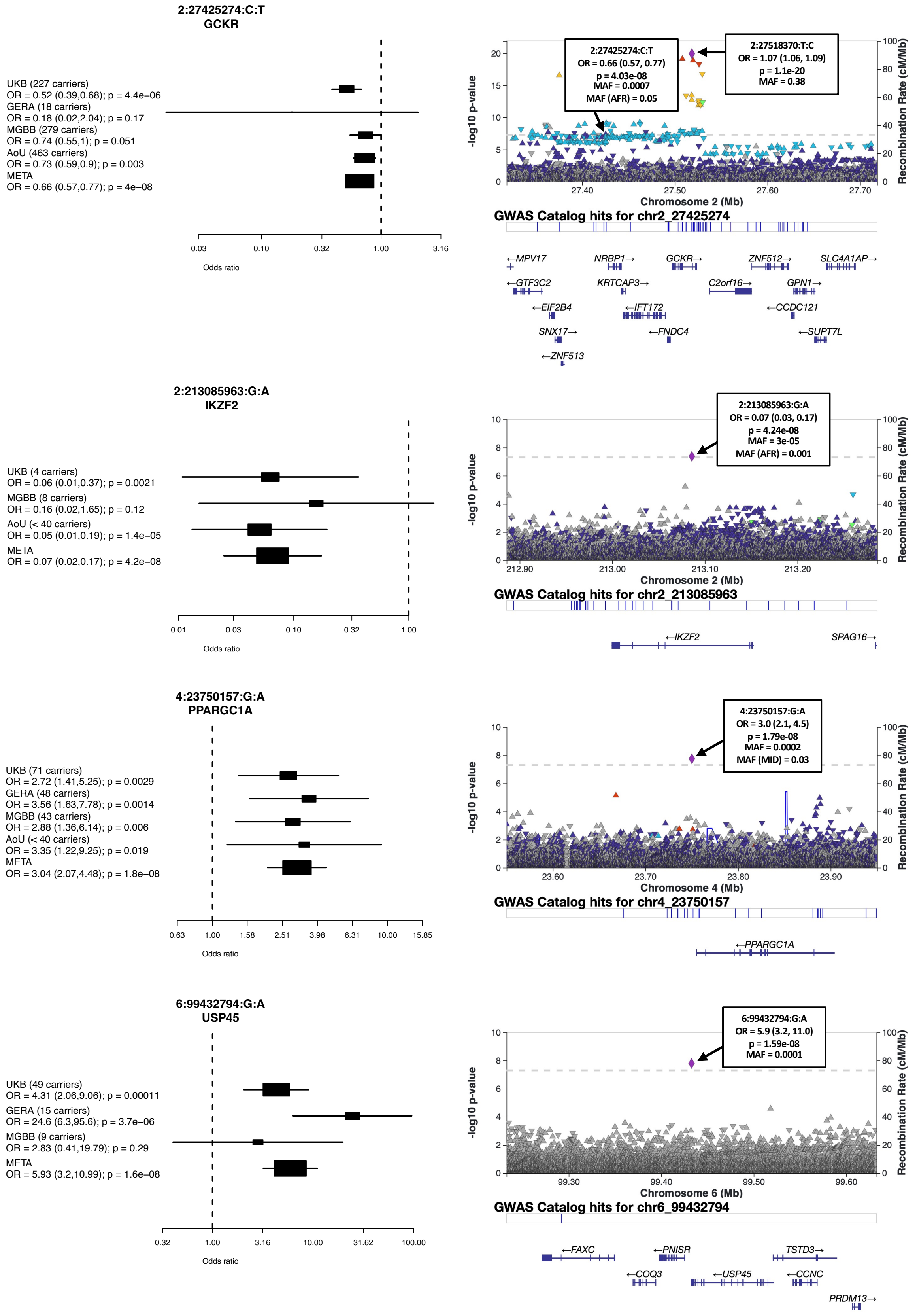

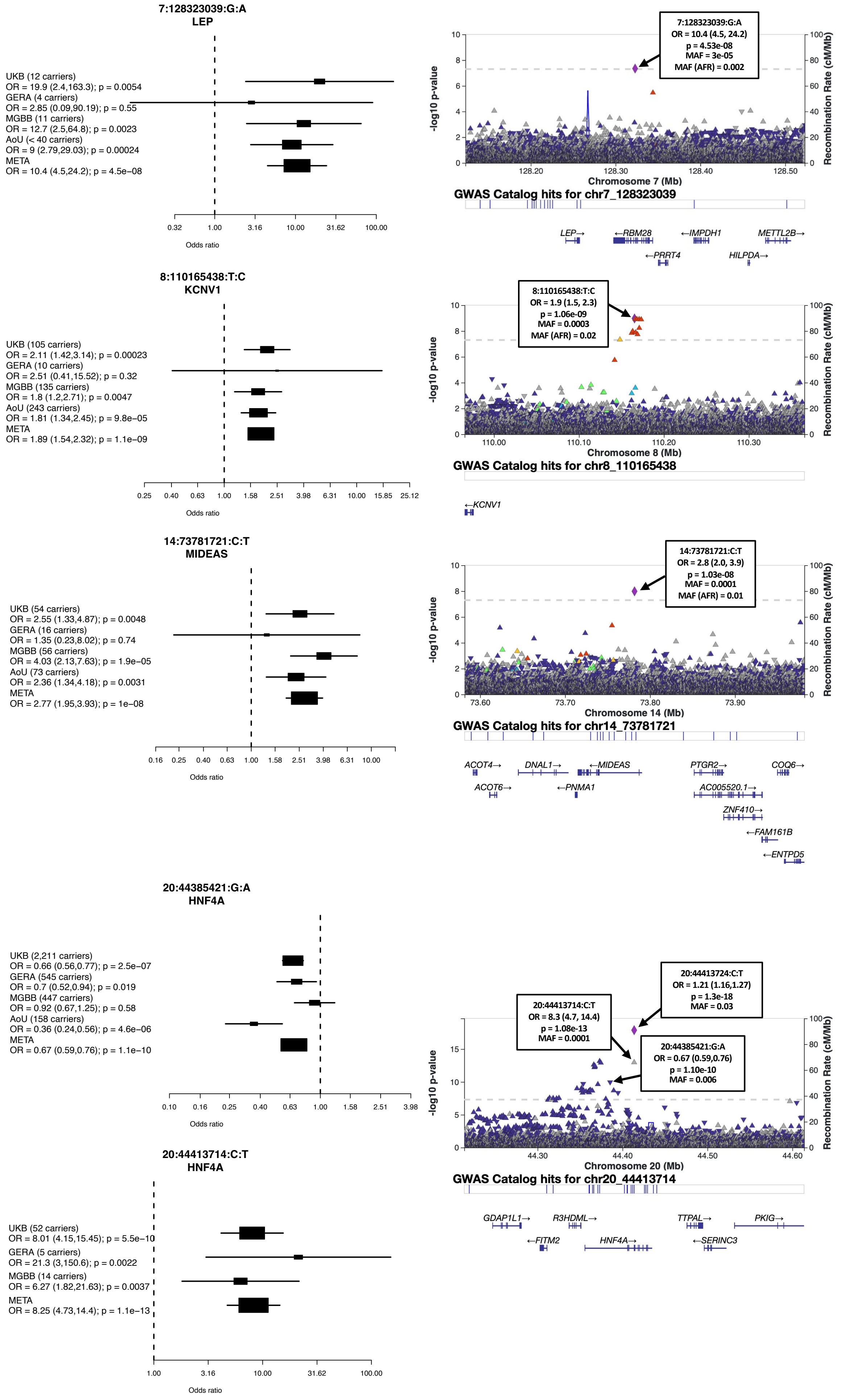

7:128323039:G:A
LEP
UKB (12 carriers)
OR = 19.9 (2.4,163.3); p = 0.0054
GERA (4 carriers)
OR = 2.85 (0.09,90.19); p = 0.55
MGBB (11 carriers)
OR = 12.7 (2.5,64.8); p = 0.0023
AoU (< 40 carriers)
OR = 9 (2.79,29.03); p = 0.00024
META
OR = 10.4 (4.5,24.2); p = 4.5e−08
0.32 1.00 3.16 10.00 31.62 100.00
Odds ratio
7:128323039:G:A
OR = 10.4 (4.5, 24.2)
p = 4.53e-08
MAF = 3e-05
MAF (AFR) = 0.002
-log10 p-value
Recombination Rate (cM/Mb)
Chromosome 7 (Mb)
GWAS Catalog hits for chr7_128323039
LEP→
←RBM28
←IMPDH1
METTL2B→
←PRRT4
HILPDA→
8:110165438:T:C
KCNV1
UKB (105 carriers)
OR = 2.11 (1.42,3.14); p = 0.00023
GERA (10 carriers)
OR = 2.51 (0.41,15.52); p = 0.32
MGBB (135 carriers)
OR = 1.8 (1.2,2.71); p = 0.0047
AoU (243 carriers)
OR = 1.81 (1.34,2.45); p = 9.8e−05
META
OR = 1.89 (1.54,2.32); p = 1.1e−09
0.25 0.40 0.63 1.00 1.58 2.51 3.98 6.31 10.00 15.85 25.12
Odds ratio
8:110165438:T:C
OR = 1.9 (1.5, 2.3)
p = 1.06e-09
MAF = 0.0003
MAF (AFR) = 0.02
Chromosome 8 (Mb)
GWAS Catalog hits for chr8_110165438
←KCNV1
14:73781721:C:T
MIDEAS
UKB (54 carriers)
OR = 2.55 (1.33,4.87); p = 0.0048
GERA (16 carriers)
OR = 1.35 (0.23,8.02); p = 0.74
MGBB (56 carriers)
OR = 4.03 (2.13,7.63); p = 1.9e−05
AoU (73 carriers)
OR = 2.36 (1.34,4.18); p = 0.0031
META
OR = 2.77 (1.95,3.93); p = 1e−08
0.16 0.25 0.40 0.63 1.00 1.58 2.51 3.98 6.31 10.00
Odds ratio
14:73781721:C:T
OR = 2.8 (2.0, 3.9)
p = 1.03e-08
MAF = 0.0001
MAF (AFR) = 0.01
Chromosome 14 (Mb)
GWAS Catalog hits for chr14_73781721
ACOT4→
DNAL1→
←MIDEAS
PTGR2→
COQ6→
ACOT6→
←PNMA1
AC005520.1→
ZNF410→
←FAM161B
←ENTPD5
20:44385421:G:A
HNF4A
UKB (2,211 carriers)
OR = 0.66 (0.56,0.77); p = 2.5e−07
GERA (545 carriers)
OR = 0.7 (0.52,0.94); p = 0.019
MGBB (447 carriers)
OR = 0.92 (0.67,1.25); p = 0.58
AoU (158 carriers)
OR = 0.36 (0.24,0.56); p = 4.6e−06
META
OR = 0.67 (0.59,0.76); p = 1.1e−10
0.10 0.16 0.25 0.40 0.63 1.00 1.58 2.51 3.98
Odds ratio
20:44413714:C:T
HNF4A
UKB (52 carriers)
OR = 8.01 (4.15,15.45); p = 5.5e−10
GERA (5 carriers)
OR = 21.3 (3,150.6); p = 0.0022
MGBB (14 carriers)
OR = 6.27 (1.82,21.63); p = 0.0037
META
OR = 8.25 (4.73,14.4); p = 1.1e−13
1.00 3.16 10.00 31.62 100.00
Odds ratio
20:44413724:C:T
OR = 1.21 (1.16,1.27)
p = 1.3e-18
MAF = 0.03
20:44413714:C:T
OR = 8.3 (4.7, 14.4)
p = 1.08e-13
MAF = 0.0001
20:44385421:G:A
OR = 0.67 (0.59,0.76)
p = 1.10e-10
MAF = 0.006
Chromosome 20 (Mb)
GWAS Catalog hits for chr20_44413714
GDAP1L1→
R3HDML→
TTPAL→
PKIG→
←FITM2
HNF4A→
←SERINC3

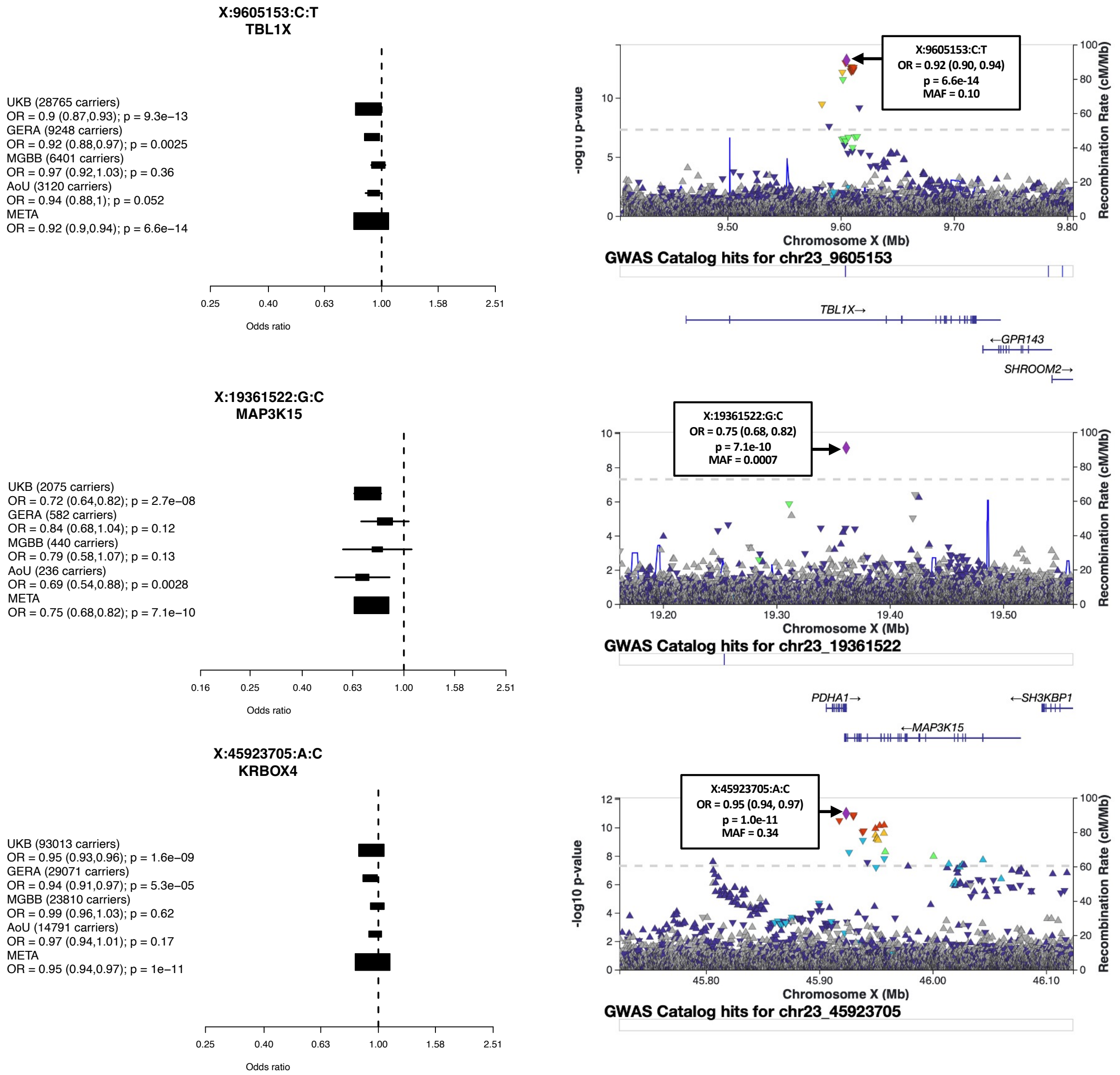

X:9605153:C:T
TBL1X
UKB (28765 carriers)
OR = 0.9 (0.87,0.93); p = 9.3e−13
GERA (9248 carriers)
OR = 0.92 (0.88,0.97); p = 0.0025
MGBB (6401 carriers)
OR = 0.97 (0.92,1.03); p = 0.36
AoU (3120 carriers)
OR = 0.94 (0.88,1); p = 0.052
META
OR = 0.92 (0.9,0.94); p = 6.6e−14
0.25
0.40
0.63
1.00
1.58
2.51
Odds ratio
X:9605153:C:T
OR = 0.92 (0.90, 0.94)
p = 6.6e-14
MAF = 0.10
-log10 p-value
Recombination Rate (cM/Mb)
Chromosome X (Mb)
GWAS Catalog hits for chr23_9605153
TBL1X→
←GPR143
SHROOM2→
X:19361522:G:C
MAP3K15
UKB (2075 carriers)
OR = 0.72 (0.64,0.82); p = 2.7e−08
GERA (582 carriers)
OR = 0.84 (0.68,1.04); p = 0.12
MGBB (440 carriers)
OR = 0.79 (0.58,1.07); p = 0.13
AoU (236 carriers)
OR = 0.69 (0.54,0.88); p = 0.0028
META
OR = 0.75 (0.68,0.82); p = 7.1e−10
0.16
0.25
0.40
0.63
1.00
1.58
2.51
Odds ratio
X:19361522:G:C
OR = 0.75 (0.68, 0.82)
p = 7.1e-10
MAF = 0.0007
Chromosome X (Mb)
Recombination Rate (cM/Mb)
GWAS Catalog hits for chr23_19361522
PDHA1→
←SH3KBP1
←MAP3K15
X:45923705:A:C
KRBOX4
UKB (93013 carriers)
OR = 0.95 (0.93,0.96); p = 1.6e−09
GERA (29071 carriers)
OR = 0.94 (0.91,0.97); p = 5.3e−05
MGBB (23810 carriers)
OR = 0.99 (0.96,1.03); p = 0.62
AoU (14791 carriers)
OR = 0.97 (0.94,1.01); p = 0.17
META
OR = 0.95 (0.94,0.97); p = 1e−11
0.25
0.40
0.63
1.00
1.58
2.51
Odds ratio
X:45923705:A:C
OR = 0.95 (0.94, 0.97)
p = 1.0e-11
MAF = 0.34
-log10 p-value
Recombination Rate (cM/Mb)
Chromosome X (Mb)
GWAS Catalog hits for chr23_45923705

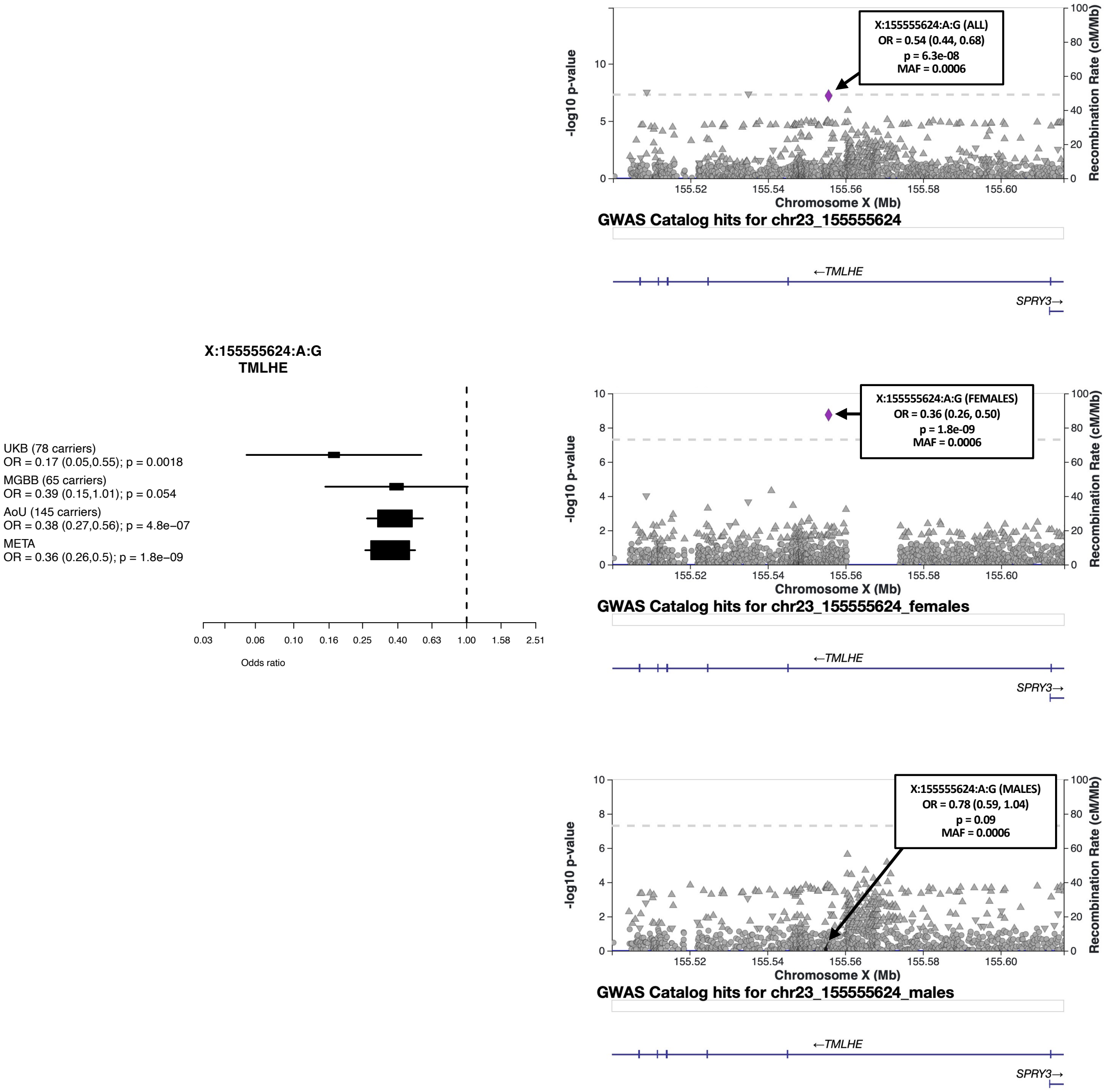
X:155555624:A:G
TMLHE
UKB (78 carriers)
OR = 0.17 (0.05,0.55); p = 0.0018
MGBB (65 carriers)
OR = 0.39 (0.15,1.01); p = 0.054
AoU (145 carriers)
OR = 0.38 (0.27,0.56); p = 4.8e−07
META
OR = 0.36 (0.26,0.5); p = 1.8e−09
0.03 0.06 0.10 0.16 0.25 0.40 0.63 1.00 1.58 2.51
Odds ratio
X:155555624:A:G (ALL)
OR = 0.54 (0.44, 0.68)
p = 6.3e-08
MAF = 0.0006
-log10 p-value
Recombination Rate (cM/Mb)
Chromosome X (Mb)
155.52 155.54 155.56 155.58 155.60
GWAS Catalog hits for chr23_155555624
←TMLHE
SPRY3→
X:155555624:A:G (FEMALES)
OR = 0.36 (0.26, 0.50)
p = 1.8e-09
MAF = 0.0006
-log10 p-value
Recombination Rate (cM/Mb)
Chromosome X (Mb)
GWAS Catalog hits for chr23_155555624_females
←TMLHE
SPRY3→
X:155555624:A:G (MALES)
OR = 0.78 (0.59, 1.04)
p = 0.09
MAF = 0.0006
-log10 p-value
Recombination Rate (cM/Mb)
Chromosome X (Mb)
GWAS Catalog hits for chr23_155555624_males
←TMLHE
SPRY3→

### Supplementary Figure 6

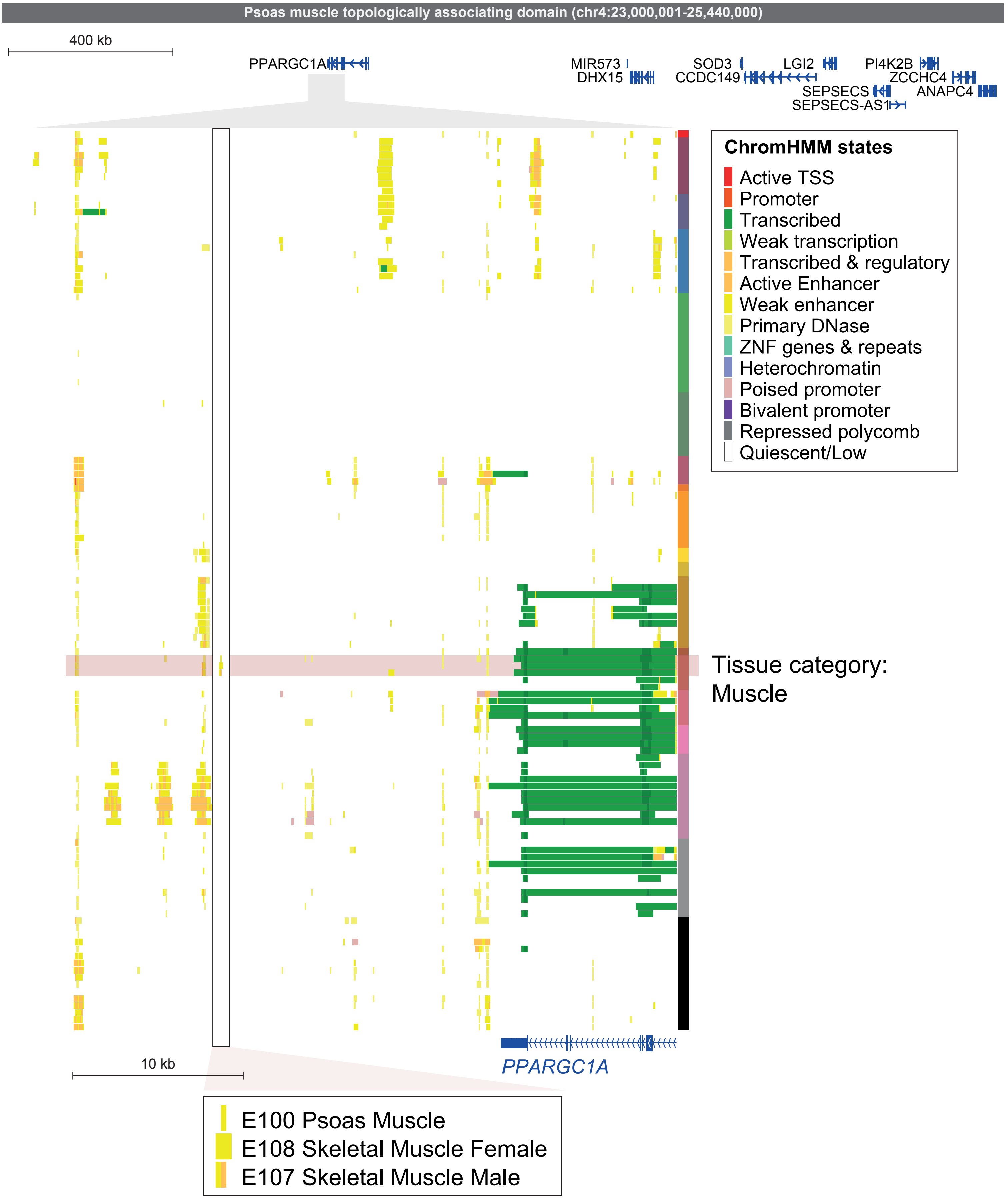

Supplementary Figure 7

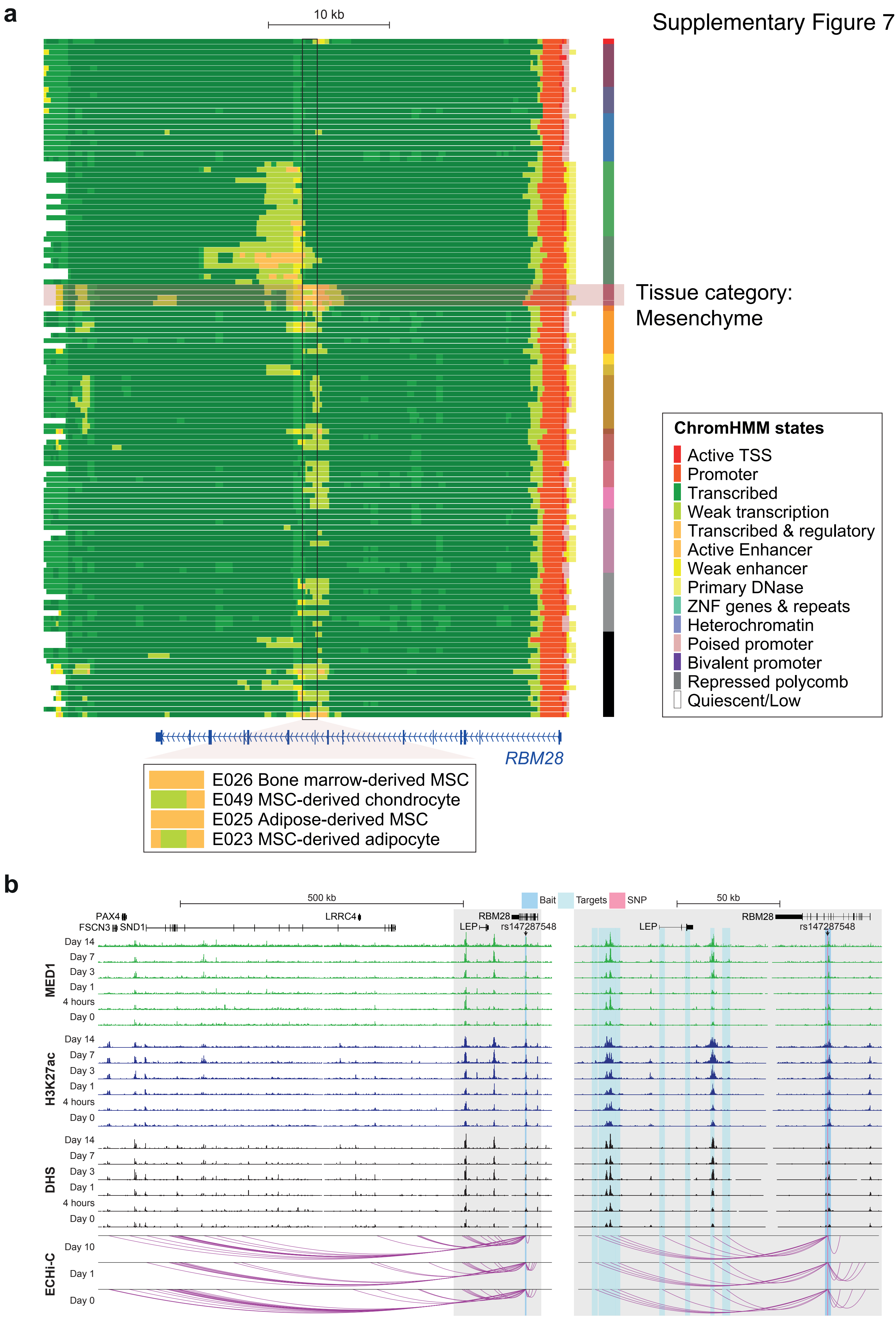

Supplementary Figure 8

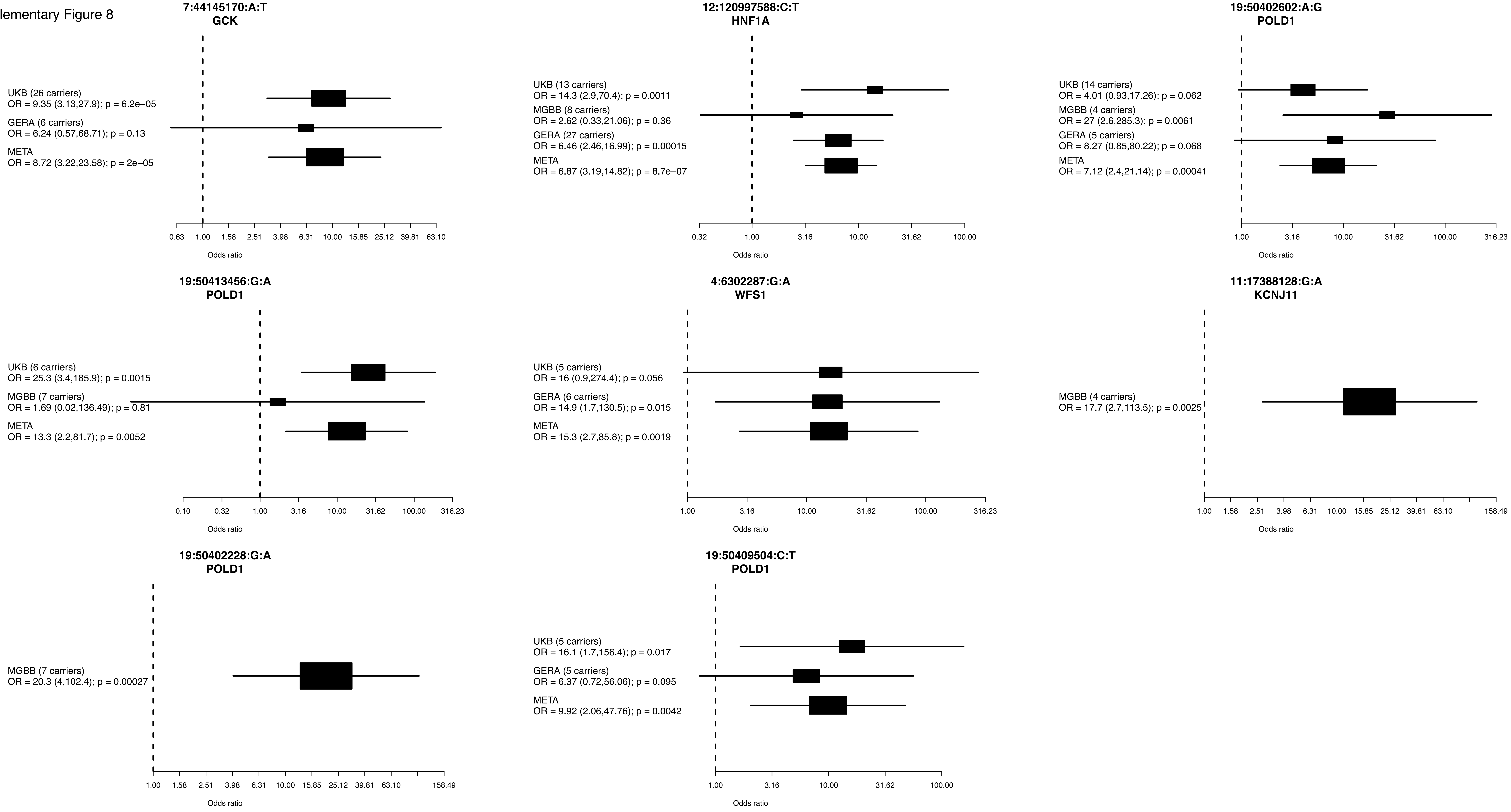

Supplementary Figure 9

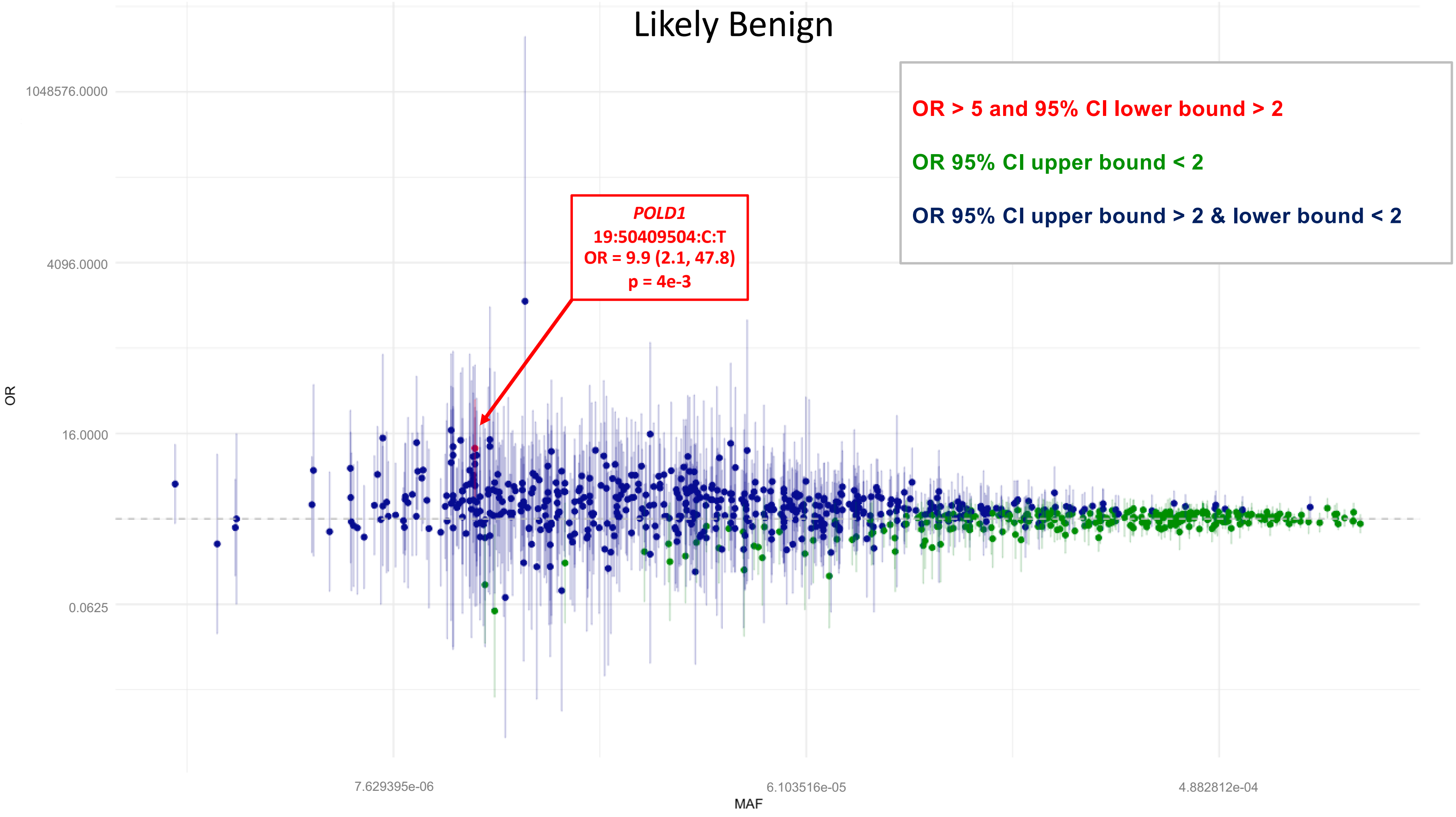

Supplementary Figure 10

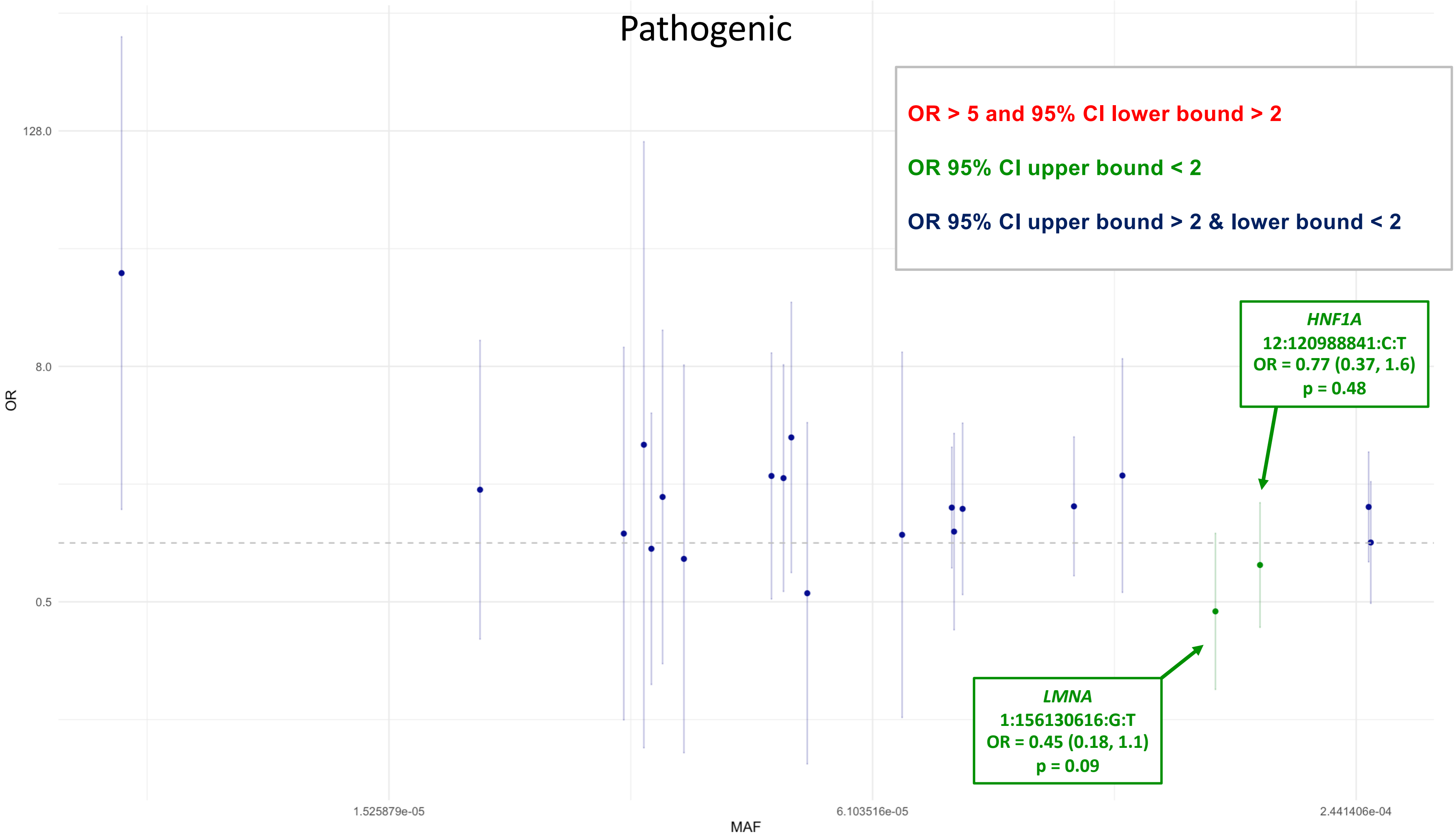

Supplementary Figure 11

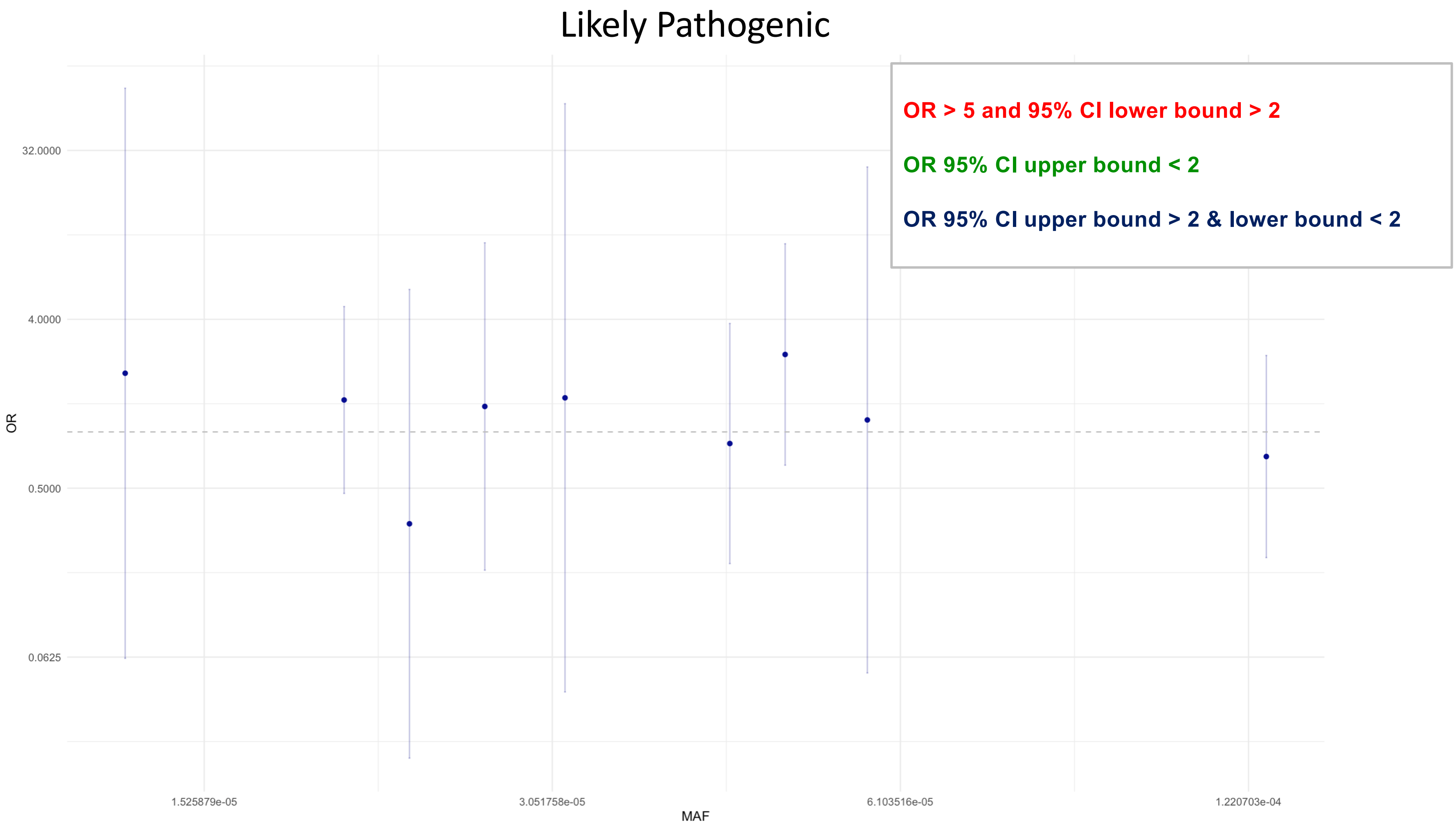
